# Role of the gut microbiome in frequent gut colonization with extended-spectrum β lactamase-producing Enterobacterales among Peruvian children

**DOI:** 10.1101/2024.11.06.24316595

**Authors:** Maya L. Nadimpalli, Neha Sehgal, Luismarcelo Rojas-Salvatierra, Robert H. Gilman, Ashlee M. Earl, Colin J. Worby, Madison Schwab, Amy J. Pickering, Monica J. Pajuelo

## Abstract

Gut colonization with extended-spectrum beta lactamase-producing Enterobacterales (ESBL-E) is increasingly common among children in low and middle-income countries. Some children nevertheless remain never or rarely colonized during early life. Understanding how this protection is conferred could be helpful for designing future interventions to protect children’s health. Here, we investigated whether differences in gut microbiome development could underlie differential susceptibility to ESBL-E gut colonization over time among children in peri-urban Lima. Weekly stool and daily surveys were collected from 345 children <3 years old during a 2016-19 study of enteric infections. A subset of children (n=12) were rarely gut-colonized with ESBL-E from 1-16 months of age. We performed short-read metagenomic sequencing of stool collected at 3, 6, 7, 9, 12, and 16 months from these children and a random subset of 42 frequently colonized children, and characterized differences in their exposures and gut microbiomes. No differences in gut taxa or functional pathways were identified over time, though children harbored differentially abundant taxa, more unique *E. coli* strains, and a higher abundance of *bla*_CTX-M_ gene copies at ESBL-E-positive versus negative timepoints. Differing patterns of ESBL-E colonization over time among children in peri-urban Lima do not appear to be related to differences in gut microbiome development.

## Introduction

Antimicrobial-resistant (AMR) pathogens are a public health threat, particularly in low- and middle-income (LMIC) countries like Peru where the infectious disease burden remains high. Gut colonization with extended-spectrum β-lactamase-producing Enterobacterales (ESBL-E), which are resistant to 3^rd^ generation cephalosporins, is common among children living in LMIC cities and is a risk factor for infections, particularly in healthcare settings.^1–5^ Recent studies of community-dwelling children and adults in LMICs in South America, South Asia, and Africa indicate that ESBL-E gut colonization rates vary widely, from 30-90%.^6–9^

Previous studies in low-resource settings have sought to identify risk factors for children’s ESBL-E acquisition.^10–12^ This is challenging to investigate in settings like peri-urban Lima, where pediatric ESBL-E gut colonization now exceeds >70%, suggesting ubiquitous environmental exposures. Some children in this setting nevertheless remain rarely or never colonized with ESBL-E early in life. Intestinal “colonization resistance” refers to the ability of gut microbiota to inhibit colonization and overgrowth of exogenous bacteria, e.g. by limiting the availability of micronutrients and carbon sources, through direct competition for adhesion receptors on the gut epithelium, and by producing toxins and antimicrobial compounds.^13^ Given that ESBL-E gut colonization is a risk factor for infection and spread, especially at higher abundances,^14,15^ identifying microbial taxa that inhibit children’s gut colonization with ESBL-E over time, as well as early-life exposures that support the assembly and maintenance of these protective taxa, could be helpful in designing future interventions to protect children’s health.

Our team recently led a prospective birth cohort study of enteric infections among 345 children living in a shantytown of peri-urban Lima. We previously used culture-based methods to characterize children’s gut colonization with ESBL-producing *Escherichia coli* and ESBL-producing *Klebsiella* spp., *Enterobacter* spp., and *Citrobacter* spp. (KEC) at or near 1, 3, 4, 5, 6, 7, 9, 12, and 16 months of age for a subset of 112 children enrolled in the cohort study.^4^ We noted that while most children were frequently gut-colonized with ESBL-E over time, a fraction (n=12) were rarely or never colonized. Here, we investigated whether differences in gut microbiome development could underlie differing patterns of susceptibility to ESBL-E gut-colonization among this population.

## Materials and methods

### Stool collection

Stool samples were collected during a prospective cohort study of childhood enteric infections in Villa El Salvador, Lima, Peru (NIH R01AI108695-01A1).^4^ Pregnant women were identified through an initial population screening in the community and through community health centers. Full-term babies were enrolled no later than 4 weeks after birth from February 2016-May 2017 (n=345), and daily surveillance data were collected through at least two years of age. Exclusion criteria for newborns participating in the parent cohort included low birth weight and presence of any severe disease requiring hospitalization early in life. Starting at recruitment and weekly thereafter, caregivers also kept a freshly soiled diaper for field worker collection; field workers retrieved these once caretakers alerted them that soiled diapers were available for pick-up. Upon arrival at a local lab facility, stool was aliquoted and stored at -20°C for up to 48h, then transported to UPCH for long-term storage at -80°C.

### Sample selection

As previously described,^4^ Enterobacterales were cultured from weekly stool samples of all newborns enrolled after December 2016 (112/345), in support of a secondary study focused on enterotoxigenic *E. coli*.^16^ For each sample, Enterobacterales colonies were pooled then archived. Colonies collected at approximately 1, 3, 4, 5, 6, 7, 9, 12, and 16 months of age were later screened for ESBL production using the double disk diffusion assay. We observed two distinct patterns: children who were never or rarely colonized with ESBL-E (n=12), hereafter “*rarely colonized*,” and children who were often or always colonized (n=100), hereafter “*frequently colonized”* (**Figures S1 and S2**). Rarely colonized children first tested positive for ESBL-E at approximately 6 months or later, and additionally were colonized at fewer than half of all tested timepoints (up to 9 per child). Frequently colonized children were colonized at more than half of all tested timepoints.

### Exposure assessment

As previously described,^4,16^ socioeconomic variables were collected at recruitment and then once yearly. Additionally, field staff visited infants every day except Sunday to collect survey data from the previous 24 hours. If field workers were unable to visit an infants’ home (e.g., day was Sunday), field workers collected survey data for the missing day at the next visit. At each daily visit, caretakers were asked about children’s milk consumption, antibiotic use, diarrhea (defined as ≥3 liquid or semi-liquid stools), and symptoms of infection (e.g. stomach pain, fever, vomiting, swelling, and loss of appetite) in the past 24 hours. Caretakers were asked to provide medication packaging so trained field workers could confirm it was an antibiotic. Because survey data were collected daily, this packaging was typically available. A new antibiotic course began when caretakers reported antibiotic consumption following two days of no exposure; a course ended when the child did not consume antibiotics for two consecutive days.

### Metagenomic sequencing

We selected stool specimens collected at approximately 3, 6, 7, 9, 12, and 16 months of age for analysis, as these were timepoints for which we had previously characterized children’s ESBL-E gut colonization, and because these timepoints span both stages of early microbiome development as previously defined by the TEDDY study,^17^ i.e., the developmental phase (3-14 months) and the transitional phase (15-30 months). We performed short-read metagenomic sequencing on available frozen stool specimens collected at these timepoints from all rarely colonized children (n=12 children, n=71 stool samples), and at these same timepoints from a randomly selected subset of 42 frequently colonized children (n=238 stool samples). We used the QIAamp PowerFecal Pro DNA Kit (Qiagen) to extract total DNA from approximately 0.25 g of frozen feces at UPCH. Extraction blanks were included with each batch of extractions. DNA was quantified using a Qubit 4 fluorometer (Invitrogen); DNA concentrations were below the Qubit level of detection (<0.1 ng/µL) for all extraction blanks. DNA extracts meeting the minimum concentration for sequencing (>10 ng/µL) were sent to the Broad Institute for short-read, paired-end 150 bp sequencing on an Illumina Novaseq 6000 System using SP4 flow cells to achieve 6 Gb per sample.

### Processing metagenomic data

Sequencing adaptors and low quality reads were removed using bbmap.^18^ Default settings were used for adaptor removal (ktrim=r k=23 mink=11 hdist=1); we retained reads with a minimum length of 50 bp (minlen=50) and a quality value of at least 20 (trimq=20). After tabulating the number of raw reads per sample, we excluded samples with more or less reads than two standard deviations from the mean from further analysis (n=11). Reads that mapped to a non-redundant version of the Genome Reference Consortium Human Build 38 (GRCh38; www.ncbi.nlm.nih.gov) using Bowtie2 v2.5.1 were considered “human contaminant sequences” and discarded. Reads with human contaminant sequences removed were deposited in the Sequence Read Archive (https://www.ncbi.nlm.nih.gov/sra) under project number PRJNA1138246 (**Table S1**).

### Profiling taxonomy, functional potential, and antibiotic resistance genes

Taxonomic assignment of short reads was performed using MetaPhlAn3 (with db v31).^19^ MetaPhlAn3 uses clade-specific marker genes to assign taxonomy at the species level; results are then aggregated to higher taxonomic levels. Alpha diversity metrics were derived from the MetaPhlAn3 relative abundance output using the vegan package in R, using the *diversity* and *richness* functions [https://cran.r-project.org/web/packages/vegan/].

Functional annotations were performed using Humann3,^20^ which uses MetaPhlAn3 output to generate organism-specific gene family and pathway abundance profiles. Briefly, for each sample, reads were aligned to a sample-specific species-level pangenome using Bowtie2.^21^ Unclassified reads were then aligned to UniRef90, a comprehensive and non-redundant protein database,^22^ using DIAMOND.^23^ Gene family abundance was annotated using UniRef90 definitions. Gene families annotated to metabolic reactions were then further analyzed to identify a parsimonious set of pathways explaining these observed reactions, using MinPath and Metacyc pathway definitions.^24,25^ We considered community-level (rather than organism-specific) metabolic pathway abundance in our statistical analyses.

Antibiotic resistance genes (ARGs) were identified by mapping short reads to the Resfinder database (v. 3.1.1) using the KMA tool.^26^ Matches with >90% coverage and >95% identity were considered true hits. We identified the number of genome equivalents in each sample using Microbe Census.^27^ To normalize for the number of bacterial genomes in each sample, ARG abundance was calculated as fragments per kilobase per million mapped reads (FPKM) divided by the total number of genome equivalents in that sample.^28^ FPKM is a common approach for comparing ARG abundance across samples, and is used to account for differences in sequencing depth between samples as well as the fact that ARGs with greater length are otherwise likely to be overrepresented, regardless of sequencing depth.^28,29^ We chose to normalize by genome equivalents, rather than total reads classified as bacteria by MetaPhlAn3, because bacterial genome sizes can vary widely. The abundance of *bla*_CTX-M_ genes (all allelic variants collectively considered as one group) and total ARGs in each sample was determined using this approach. FPKMs were log_10_ transformed for analysis.

### Characterizing *E. coli, Klebsiella* spp., and Enterobacter *spp.* strain diversity

We used the StrainGE tool^30^ to characterize *E. coli, Klebsiella* spp., and Enterobacter *spp.* strains in children’s fecal metagenomes. We theorized that commensal strains could play a role in excluding ESBL-producing *E. coli* and KEC from the gut environment through species-specific effects. StrainGE can sensitively profile strains in a microbial community with >0.1% relative abundance.^30^ A reference database was built using complete *E. coli* genomes from NCBI’s RefSeq database (3,108 genomes downloaded March 5, 2024), and each genome was assigned a Clermont phylotype using EzClermont v.0.70.^31^ A separate reference database was built using complete *Klebsiella* spp. and *Enterobacter* spp. genomes from NCBI’s RefSeq database (4,121 genomes downloaded June 5, 2025). *Citrobacter* spp., were relatively rare (0.014% abundance on average) and therefore were not considered. Briefly, references in each database were kmerized using the *straingst* kmerize command and the default k-mer size of 23. To reduce redundancy, closely related genomes were clustered based on k-mer similarity, using the *straingst kmersim* and *straingst cluster* commands. A single pangenome k-mer database was then created for *E. coli* and for *Klebsiella* spp./*Enterobacter* spp., respectively, using *straingst createdb.* To align sample metagenome reads to each pangenome k-mer database, reads were first kmerized using *straingst kmerize*. We then used *straingst run* to identify reference strains present in each sample.

### Data analysis

We examined differences in sociodemographic characteristics, diarrhea rates, and antibiotic use between rarely and frequently gut-colonized children using t-tests and chi square tests for all 112 children for whom we characterized ESBL-E gut colonization, and separately for the 54 children whose stool samples were sequenced.

For the subset of 54 children with fecal metagenomes, we examined whether any taxa or metabolic pathways were differentially abundant between rarely and frequently gut-colonized children across time using mixed effects linear regression models. An arcsine square root transformation was applied to relative abundance values. We also used mixed effects linear regression models to examine differences in log10 transformed species richness values, Shannon diversity, *bla*_CTX-M_ FPKM and total ARG FPKM over time between groups. Transformed relative abundance values, alpha diversity metrics, and ARG FPKM were modeled as a function of subject (random effects term), colonization pattern, i.e., rarely versus frequently colonized (binary fixed effects term), child age (continuous fixed effects term), and hours before diaper retrieval (continuous fixed effects term). We also ran additionally parametrized models adjusted for factors known to affect the gut microbiome, including child’s delivery mode (binary fixed effects term) and number of diarrhea episodes and antibiotic courses in the past 30 days (continuous fixed effects terms), as well as socioeconomic characteristics that differed between groups with p<0.20 (*i.e.*, maternal education and presence of a flush toilet; availability of piped water was not considered as it was highly collinear with presence of a flush toilet). We did not adjust for breastfeeding as there was minimal variation in this exposure over time (**Table S2**). All taxa or metabolic pathways detected in at least 10% of samples were fitted using the nlme package in R. Significance of associations was determined using Wald’s test, and p-values were adjusted for multiple hypothesis testing using the Benjamini-Hochberg correction.

For the subset of metagenomes for which we had matched ESBL-E gut colonization status, we examined whether any taxa or metabolic pathways were differentially abundant between timepoints where children were ESBL-E positive versus negative using mixed effects logistic regression models. For any given taxa or metabolic pathway detected in at least 10% of samples, we modeled ESBL-E positivity (yes/no) as a function of subject, child age, and hours before diaper retrieval and the transformed relative abundance value of that taxa. We ran additionally parametrized models that adjusted for number of recent diarrhea episodes and antibiotic courses, maternal education, and presence of a flush toilet. We used the lme4 package in R and adjusted p-values using the Benjamini-Hochberg correction. For this same subset of metagenomes, we examined whether log10 transformed species richness values, Shannon diversity, *bla*_CTX-M_ genes or total ARGs differed between timepoints where children were ESBL-E positive versus negative, using similar model specifications as above.

We compared the number of unique *E. coli, Klebsiella* spp., and *Enterobacter* spp. strains between rarely and frequently colonized children using Welch two sample t-tests, which do not assume equal variance between groups. We used Welch two sample t-tests to compare strain richness between groups overall (pooling all timepoints together), and at specific timepoints. Similarly, we compared strain richness at timepoints where children were ESBL-E positive versus negative.

Statistical significance was defined by αL=L0.05 and p-values are two-sided unless stated otherwise. All analyses were conducted in R Studio version (2024.04.1+748).

## Results

### Exposure profiles were similar between children who were rarely versus frequently gut-colonized with ESBL-E

There were no significant differences in exposures that are well-known to impact the gut microbiome, including rates of vaginal birth, antibiotic consumption rates, children’s age at time of first antibiotic consumption, or numbers of diarrhea episodes per child month between rarely versus frequently colonized children (**Table 1**). Rarely colonized children did consume slightly fewer antibiotic courses overall (3.4 vs 4.5 courses/child-year, *p*=0.2), and fewer azithromycin courses in particular (0.1 vs 0.5 courses/child-year, *p*=0.14), but differences were not statistically significant. Only 2/112 children did not consume any antibiotics, yet both were frequently colonized with ESBL-E.

**Table 1.**
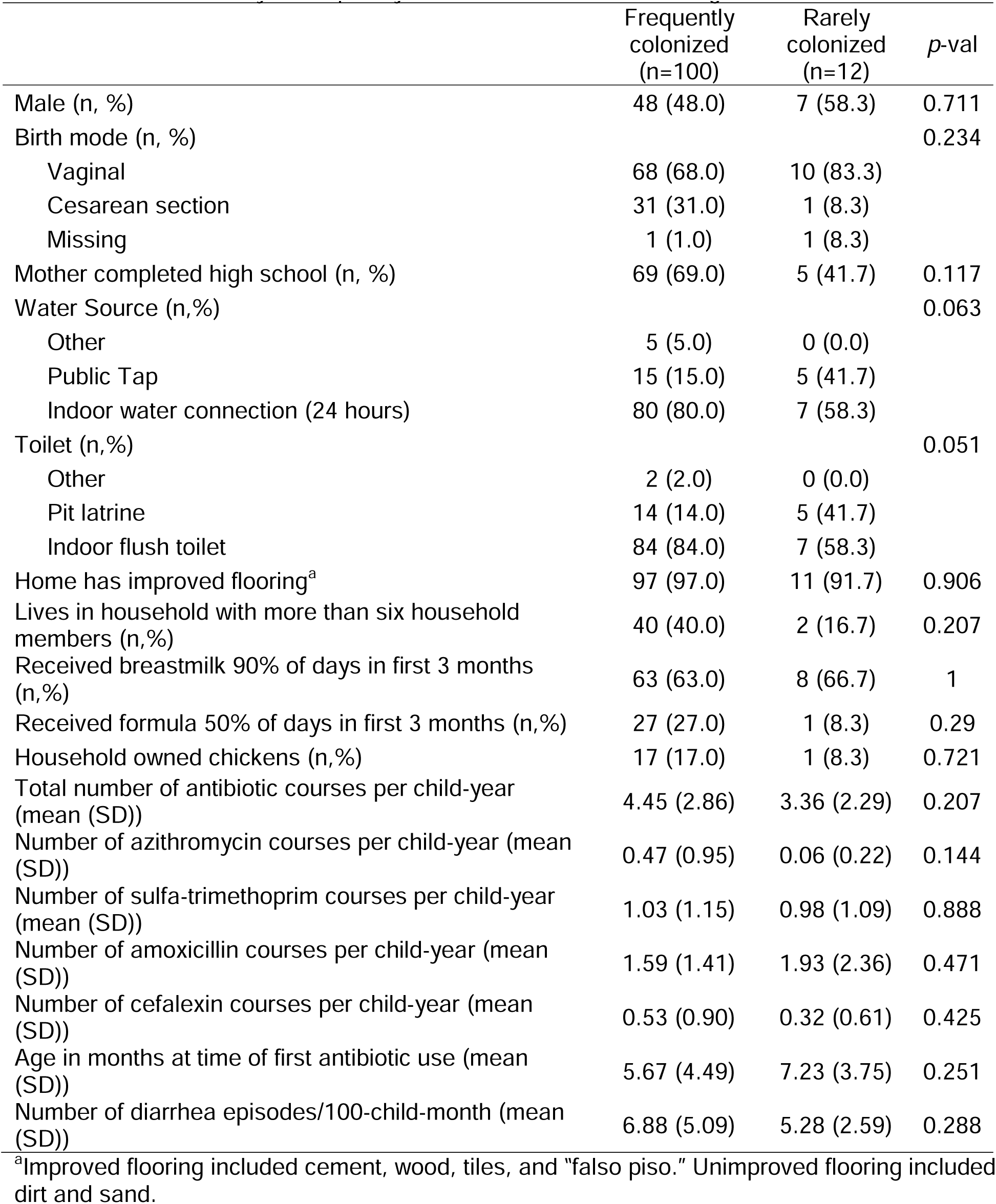
Sociodemographic characteristics and early-life exposures among 112 Peruvian children who were rarely or frequently colonized with ESBL-E during the first 16 months of life.

|  | Frequently colonized (n=100) | Rarely colonized (n=12) | p-val |
| --- | --- | --- | --- |
| Male (n, %) | 48 (48.0) | 7 (58.3) | 0.711 |
| Birth mode (n, %) |  |  | 0.234 |
| Vaginal | 68 (68.0) | 10 (83.3) |  |
| Cesarean section | 31 (31.0) | 1 (8.3) |  |
| Missing | 1 (1.0) | 1 (8.3) |  |
| Mother completed high school (n, %) | 69 (69.0) | 5 (41.7) | 0.117 |
| Water Source (n,%) |  |  | 0.063 |
| Other | 5 (5.0) | 0 (0.0) |  |
| Public Tap | 15 (15.0) | 5 (41.7) |  |
| Indoor water connection (24 hours) | 80 (80.0) | 7 (58.3) |  |
| Toilet (n,%) |  |  | 0.051 |
| Other | 2 (2.0) | 0 (0.0) |  |
| Pit latrine | 14 (14.0) | 5 (41.7) |  |
| Indoor flush toilet | 84 (84.0) | 7 (58.3) |  |
| Home has improved flooring<sup>a</sup> | 97 (97.0) | 11 (91.7) | 0.906 |
| Lives in household with more than six household members (n,%) | 40 (40.0) | 2 (16.7) | 0.207 |
| Received breastmilk 90% of days in first 3 months (n,%) | 63 (63.0) | 8 (66.7) | 1 |
| Received formula 50% of days in first 3 months (n,%) | 27 (27.0) | 1 (8.3) | 0.29 |
| Household owned chickens (n,%) | 17 (17.0) | 1 (8.3) | 0.721 |
| Total number of antibiotic courses per child-year (mean (SD)) | 4.45 (2.86) | 3.36 (2.29) | 0.207 |
| Number of azithromycin courses per child-year (mean (SD)) | 0.47 (0.95) | 0.06 (0.22) | 0.144 |
| Number of sulfa-trimethoprim courses per child-year (mean (SD)) | 1.03 (1.15) | 0.98 (1.09) | 0.888 |
| Number of amoxicillin courses per child-year (mean (SD)) | 1.59 (1.41) | 1.93 (2.36) | 0.471 |
| Number of cefalexin courses per child-year (mean (SD)) | 0.53 (0.90) | 0.32 (0.61) | 0.425 |
| Age in months at time of first antibiotic use (mean (SD)) | 5.67 (4.49) | 7.23 (3.75) | 0.251 |
| Number of diarrhea episodes/100-child-month (mean (SD)) | 6.88 (5.09) | 5.28 (2.59) | 0.288 |

<sup>a</sup>Improved flooring included cement, wood, tiles, and “falso piso.” Unimproved flooring included dirt and sand.

Among children who were rarely gut-colonized with ESBL-E, there was a positive but non-significant trend towards living in households of lower socioeconomic status (SES). Specifically, these children were somewhat more likely to live in households that relied on a public tap for drinking water (*p*=0.06) versus having an indoor connection, and more likely to rely on pit latrines versus indoor flush toilets (*p*=0.05). Most households that relied on water taps also used pit latrines, and vice versa. Rarely colonized children were also somewhat less likely to be born to a mother who had completed high school (p=0.11), although this difference was not statistically significant. Children’s antibiotic use (total courses/child-year) was weakly correlated with household reliance on public water taps (*r_pb_*=0.24, *p*<0.05 by point-biserial correlation) and use of a pit latrine (*r_pb_*=0.25, *p*<0.05), but was not correlated with maternal education attainment (*r_pb_*=0.04, *p*=0.67).

Breastfeeding was exceptionally common among all children. On average, children received breastmilk nearly every day in the 30 days prior to a stool sample through 16 months of age (**Table S2**). Notably, nearly 1/3 children who were frequently colonized with ESBL-E regularly consumed formula early in life, compared to just 1 of 12 rarely colonized children (**Table 1**). However, this difference was not significant (p=0.29).

The distributions of these exposures were similar among the subset of 54/112 children whose fecal metagenomes were sequenced for this study, as well as the 274/345 children participating in the parent cohort who completed at least 16 months of data collection (**Table S3**). Of note, both children whose ESBL status was previously ascertained (n=112) and children whose fecal metagenomes were sequenced for this study (n=54) were more likely to be born by Cesarean section than children who completed 16 months of data collection in the parent cohort study (n=274) (**Table S3**).

### Peruvian children’s gut microbiomes are dominated by *Bifidobacterium* spp. regardless of ESBL-E colonization status

Samples included in our final analysis included 66 fecal metagenomes from 12 rarely colonized children and 232 fecal metagenomes from 42 frequently colonized children (n=298 total, 4-6 samples per child).

As expected given the high proportion of breastfeeding in this setting, children’s gut microbiomes were largely dominated by *Bifidobacterium* spp., comprising 58% of bacterial reads in average at 3 months, and 34% by 16 months (**Figure 1**). *B*. *longum* and *B. breve* predominated earlier in life (comprising up to 20% and 12% of bacterial reads, respectively, until 9 months of life), while *B. bifidum* consistently comprised between 10-15% of bacterial reads (**Figure 2**). The number of unique *Bifidobacterium* spp. in children’s guts ranged from 0-9 (median=4) and increased as children aged (β=0.14, SE=0.019, *p*-value<0.0001). Pooling all timepoints together, there was no difference in the relative abundance of any *Bifidobacterium* spp. between children who were rarely versus frequently gut-colonized with ESBL-E. Likewise, the number of unique *Bifidobacterium* spp. did not differ between groups.

**Figure 1.**
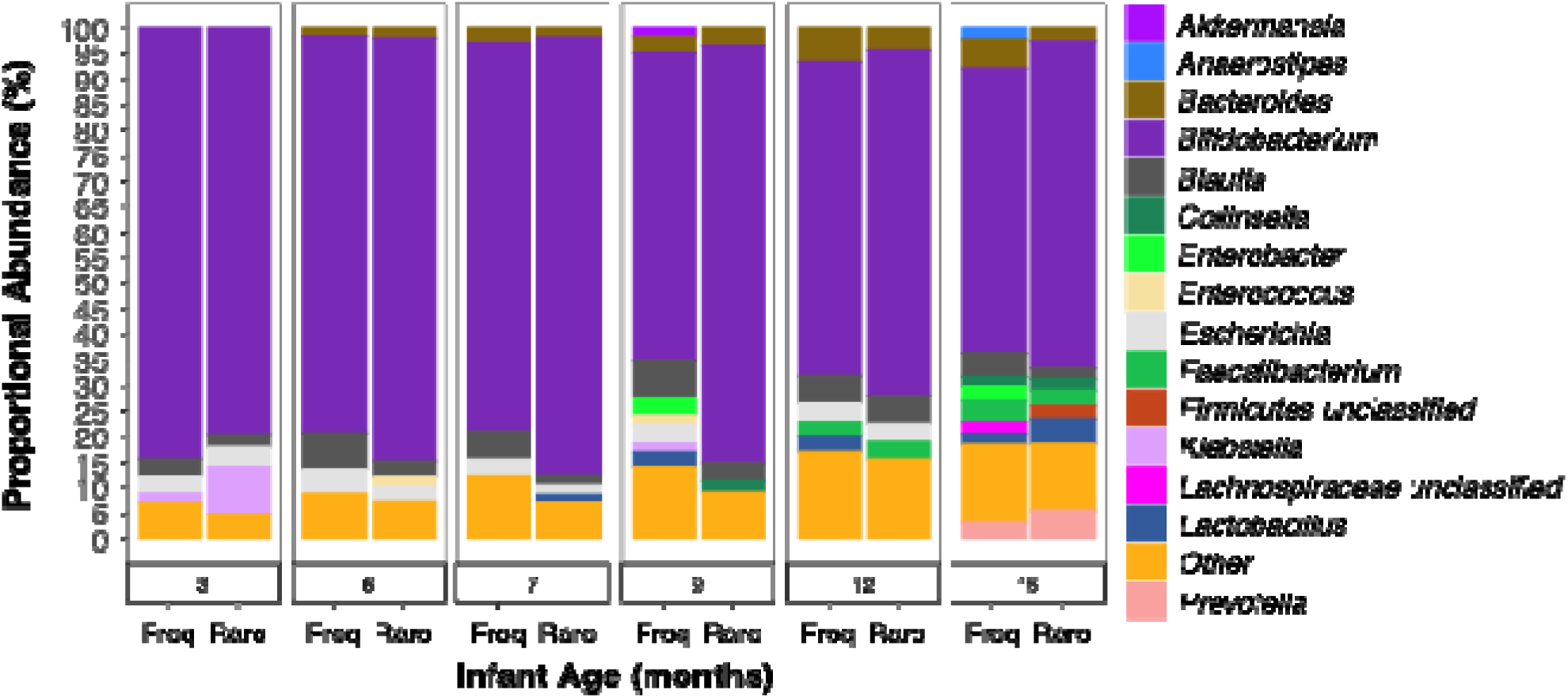
Proportion of bacterial genera with at least 1% average relative abundance in stool samples collected from Peruvian children aged 3-16 months. There were no differences in the relative abundance of any bacterial taxa between children who were rarely versus frequently gut-colonized with ESBL-E across all timepoints. Taxonomy profiling was performed using MetaPhlAn3.

**Figure 2.**
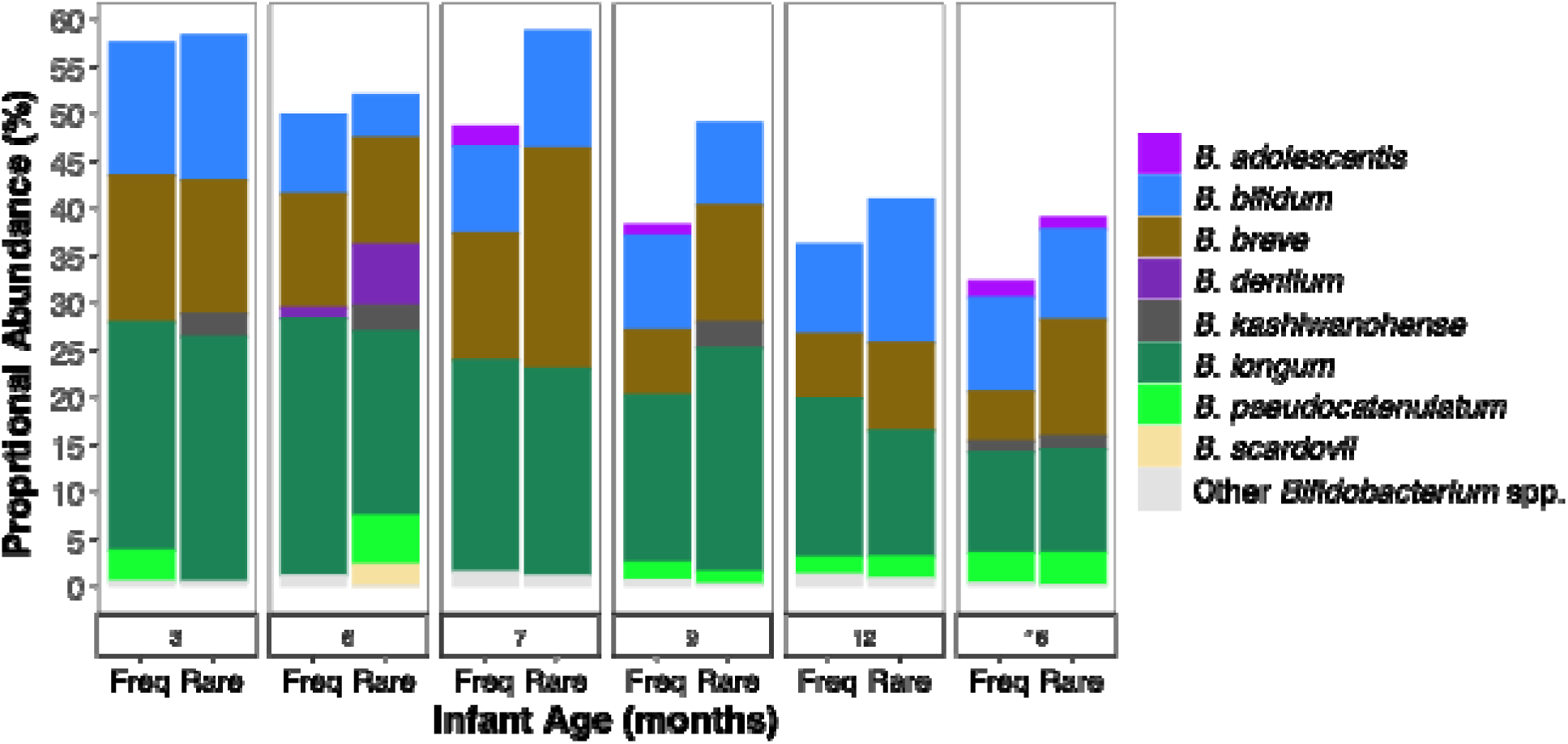
Relative abundance of dominant *Bifidobacterium spp.* in stool samples collected from Peruvian children aged 3-16 months. There were no differences in the relative abundance of any *Bifidobacterium* spp. between children who were rarely versus frequently gut-colonized with ESBL-E across all timepoints; the number of unique *Bifidobacterium* spp. also did not differ between groups. Taxonomy profiling was performed using MetaPhlAn3. *<u>Note</u>*: Fifteen *Bifidobacterium spp.* were detected among all children; species with at least 1% average relative abundance are depicted here.

### No differences in diversity, relative abundance, or functional potential of gut bacteria between rarely and frequently colonized children

Besides *Bifidobacterium*, other dominant bacterial genera included *Blautia* (average across all samples 3.1%), *Bacteroides* (2.1%), *Escherichia* (1.9%), *Lactobacillus* (1.1%), *Faecalibacterium* (0.9%), and *Klebsiella* (0.8%) (**Figure 1**). Potential enteropathogens like *Klebsiella pneumoniae* and *Enterococcus faecium* were detected among nearly all children (**Table S4**). After adjustment for multiple comparisons, no taxa were differentially abundant between children who were rarely versus frequently gut-colonized with ESBL-E over time in fully parameterized models (corrected p-values all > 0.89). Results were similar for simpler models that only adjusted for children’s colonization pattern, child age, and hours before diaper retrieval (fdr-corrected p-values all >0.66).

Like others,^17^ we found the number of unique bacterial species in children’s fecal metagenomes increased as they aged, from 31 to 75, on average. The diversity of children’s fecal metagenomes also increased (0.99 Shannon diversity index on average at 3 months, to 2.31 at 16 months) (**Figure S3**). Pooling all timepoints together, we observed no differences in diversity or richness between groups (p-values> 0.62). There were also no differences in median richness or Shannon diversity at any specific sampling point (p>0.52 for all pairwise differences by Wilcoxon rank sum tests).

A total of 449 metabolic pathways were identified in fecal metagenomes from at least 10% of children. After adjustment for multiple comparisons, no metabolic pathways were differentially abundant between children who were rarely versus frequently gut-colonized with ESBL-E over time in fully parameterized models (fdr-corrected p-values all > 0.99). Results were similar for simpler models that only adjusted for children’s colonization pattern, child age, and hours before diaper retrieval (fdr-corrected p-values all >0.87).

### Some differences in *Klebsiella* and *Enterobacter* strain richness between rarely and frequently colonized children

*E. coli* strains were detected in every sample. Among the 252 unique *E. coli* strains detected overall, most belonged to phylotypes A and B1 (**Figure 3A**). There were no differences in the number of *E. coli* strains harbored by rarely and frequently colonized children, overall or by timepoint (p>0.15 for all pairwise comparisons by Welch two-sample t-test) (**Figure 3A**). There was no difference in the proportion of strains belonging to any given *E. coli* phylotype between rarely and frequently colonized children at any time point (p>0.26 for all pairwise comparisons by Fisher test).

**Figure 3.**
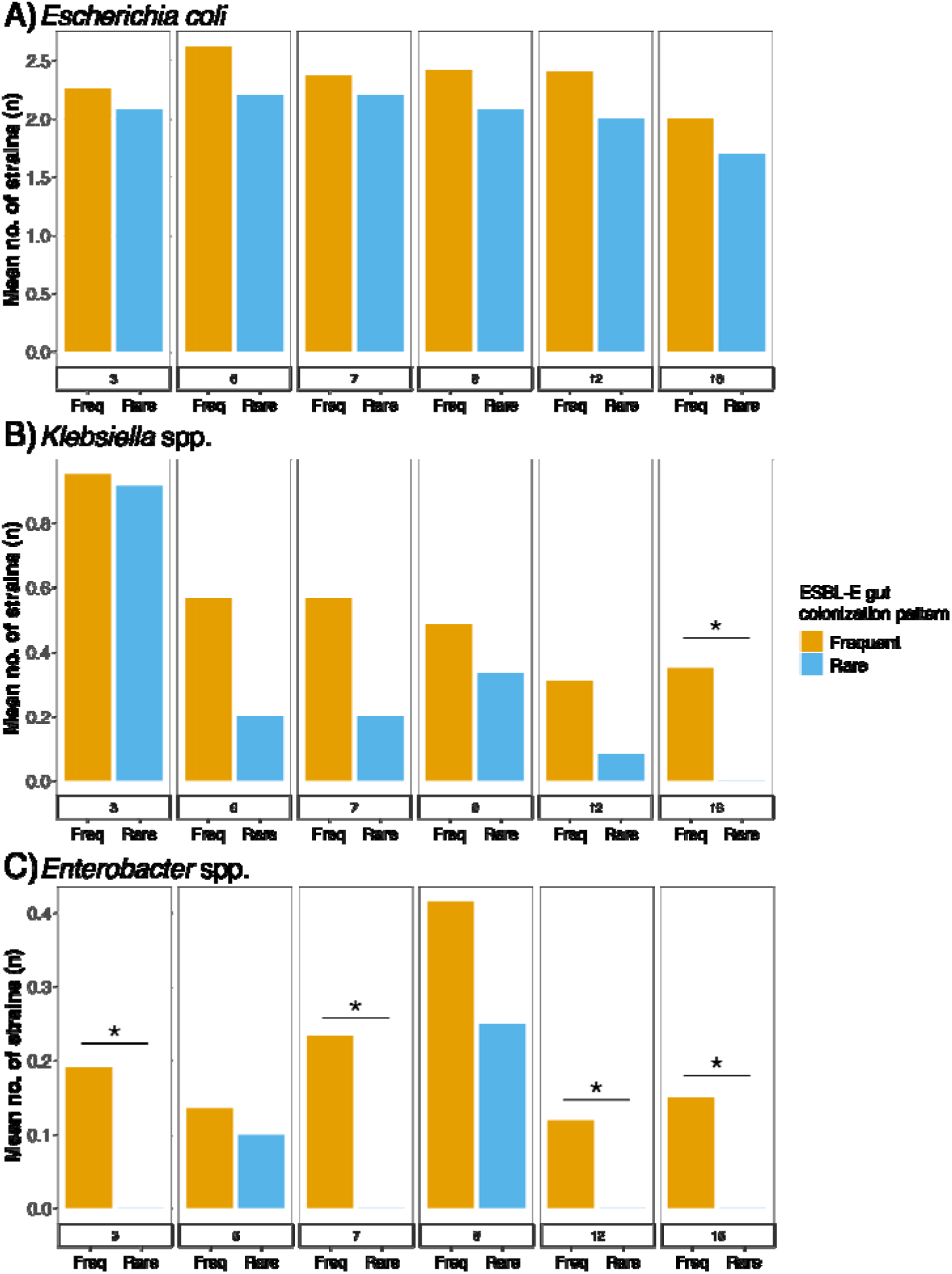
Distribution of *E. coli*, *Klebsiella* spp, and *Enterobacter* spp. strains in stool samples from Peruvian children aged 3-16 months who were frequently vs rarely gut-colonized with ESBL-E. Strains were identified using the StrainGE toolkit. Pooling all timepoints together, there were significantly fewer *Klebsiella* spp. and *Enterobacter* spp. strains detected among rarely versus frequently colonized children by Welch two-sample t-test. Note: * indicates statistically significant pairwise difference in mean number of strains by Welch two-sample t-test.

*Klebsiella* spp. strains were detected in approximately one-third of samples (106/298). Among the 70 unique strains detected overall, most were *K. pneumoniae* (66/70). Rarely colonized children harbored fewer *Klebsiella* spp. strains on average (mean=1.7), pooling all timepoints together, than frequently colonized children (mean=3.0, *p*=0.03 by Welch two-sample t-test), (**Figure 3B**). *Enterobacter* strains were rarer, detected in only 13% of samples (39/298). *E. hormaechei* (n=9), *E. asburiae* (n=4), and *E. cloacae* (n=4) comprised the 17 *Enterobacter* strains detected. On average and across all timepoints, rarely colonized children harbored fewer *Enterobacter* spp. strains (mean=0.3) than frequently colonized children (mean=1.1, *p*=0.02 by Welch two-sample t-test) (**Figure 3C**).

### Some differences in ARG abundance between rarely versus frequently colonized children

Rarely colonized children harbored a significantly lower abundance of *bla*_CTX-M_ genes (*p*<0.0001) in their stool, on average across all time points (mean log10 *bla*_CTX-M_ FPKM = 4.1, SD=4.6), compared to persistently colonized children (mean log10 *bla*_CTX-M_ FPKM = 4.9, SD=5.4) (**Figure S4**). We noted significant differences at the ages of 6 (p=0.002) and 16 months (p=0.006) by Welch two-sample test. However, there was no difference in total ARG abundance, on average across all timepoints, between these groups (among rarely colonized children: mean log10 total ARG FPKM=6.9, SD=1.9, among persistently colonized children: mean log10 total ARG FPKM=7.0, SD=7.1).

### Differences in children’s gut microbiomes and resistomes at ESBL-E positive versus negative sampling points

We also examined differences in children’s gut microbiomes and resistomes at timepoints where they were ESBL-E colonized versus not (ESBL-E gut colonization status available for 263/298 fecal metagenomes), regardless of whether they were rarely or frequently gut-colonized with ESBL-E over time.

We noted several differences in children’s gut microbiomes at timepoints when they were ESBL-E colonized. First, the relative abundance of the genus *Slackia* (0.006%, fdr-corrected p-val=0.047) in children’s stool was higher at timepoints where children were not gut-colonized with ESBL-E by culture, when controlling for child age, hours before diaper retrieval, and repeated measurements (**Figure S5**). There were also non-significant trends towards a higher abundance of *Erysipelotrichia* (0.47%; fdr-corrected p-val=0.18), *Proteobacteria* (0.03%, fdr-corrected p-val=0.16) and *Bacteroidetes* (0.03%; fdr-corrected p-val=0.16) at timepoints where were children colonized with ESBL-E by culture, and a trend towards lower abundance of *Actinobacteria* at these timepoints (46%; fdr-corrected p-val=0.12). No taxa were differentially abundant in fully parameterized models that additionally adjusted for recent diarrhea, recent antibiotic use, maternal education, and presence of a flush toilet. Second, children harbored more unique *E. coli* strains at sampling points where they were gut-colonized with ESBL-E (mean=2.3 across all age groups) versus sampling points where they were not (mean=2.1, p=0.05 by Welch two-sample t-test). However, there were no differences in the number of *Klebsiella* spp. or *Enterobacter* spp. strains harbored by children at ESBL-positive versus negative timepoints (*p*>0.25 for both comparisons). There were no differences in species richness or diversity at timepoints where children were ESBL-E colonized versus not, nor any differentially abundant metabolic pathways.

There were also significant differences in children’s gut resistomes between sampling points where children were gut-colonized with ESBL-E versus not (**Figure 4**). On average, children harbored significantly higher *bla*_CTX-M_ gene FPKM (*p*<0.0001) at sampling points where they were gut-colonized with ESBL-E (mean log10 *bla*_CTX-M_ FPKM=4.9, SD=5.4) versus sampling points where they were not (mean log10 *bla*_CTX-M_ =4.4, SD=5.2). We noted significant differences at all sampling points by Welch two-sample test (all *p*≤0.002). On average and across all timepoints, children also harbored significantly higher total ARG FPKM at sampling points (*p*=0.005) where they were gut-colonized with ESBL-E (mean log10 *bla*_CTX-M_ FPKM=7.02, SD=7.1) versus sampling points where they were not (mean log10 *bla*_CTX-M_ =6.9, SD=6.8). We noted a significant difference at 7 months of age (p=0.03) by Welch two-sample text.

**Figure 4.**
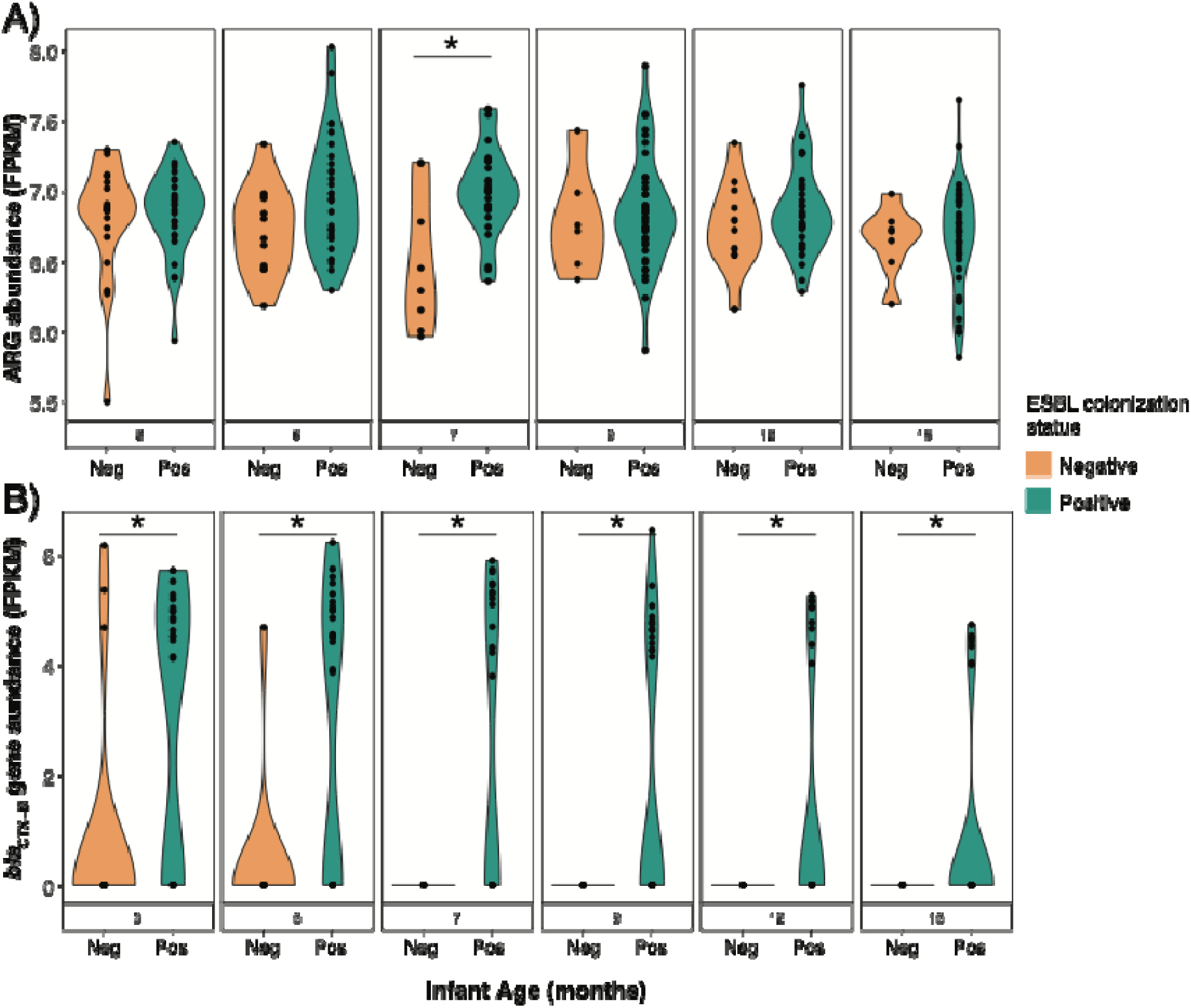
Abundance of A) total antibiotic resistance genes and B) *bla*_CTX-M_ resistance genes in Peruvian children’s stool from 3-16 months of age. The abundance of total ARGs and *bla*_CTX-M_ resistance genes were significantly higher when children were gut-colonized with ESBL-producing Enterobacterales versus not, on average and across all timepoints. *<u>Note</u>*: * denotes a statistically significant difference by Welch two sample t-test. FPKM=Fragments per kilobase per million mapped reads. FPKM values have been log transformed.

## Discussion

This is the first longitudinal study of the gut microbiome among South American children. As previously reported, gut colonization with ESBL-E was prevalent among children in this peri-urban setting and steadily increased as children aged.^4^ Similar to recent studies among healthy adults,^32–34^ our findings suggest that differences in gut microbiome development do not underlie patterns of ESBL gut colonization over time among children in peri-urban Lima, though subtle differences may have been missed due to our small sample size. To our knowledge, this is the first study to examine this research question among young children whose gut microbiomes are rapidly developing.

Three recent Western studies and two LMIC studies have investigated associations between ESBL-E gut colonization, gut microbial taxa, and/or the gut metabolome at single time points.^32–36^ Two case-control studies among Dutch^33^ and French adults^34^ found no differences in alpha diversity, taxa abundance, or microbial functional annotations between healthy adults who were gut-colonized with ESBL-E versus not, after controlling for age, sex, ethnicity, and recent travel. Another study among US travelers^32^ found no difference in the pre-travel gut microbiomes of individuals who acquired ESBL-E while abroad versus those who did not, suggesting that differences in exposures while traveling, rather than inherent differences in intestinal colonization resistance, are more important determinants of ESBL-E acquisition. One study of healthy Thai adults^36^ reported differences in gut taxa between ESBL-E carriers and non-carriers, but because potential confounders were not controlled for, it is challenging to attribute differences to ESBL-E colonization status. A small study among indigenous persons in French Guyana identified some differences in gut microbial taxa between 8 cases and 24 controls, but whether these differences preceded or were a result of ESBL-E gut colonization could not be determined.^35^ To our knowledge, no previous study has examined whether differences in the gut microbiome over time, rather than just at a single timepoint, may underlie ESBL-E gut colonization patterns. Our study adds to the existing literature by indicating that patterns of rare versus frequent ESBL-E gut colonization among children who are frequently exposed to enteropathogens and antibiotics may not primarily be a function of intestinal colonization resistance.

Taxa that previous studies have identified to be associated with ESBL carriage or non-carriage differed from what we observed. *Erysipelotrichia*, *Proteobacteria*, and *Bacteroidetes*, which are commonly isolated in children’s stool,^37,38^ were somewhat more abundant at ESBL-positive timepoints (*p-val*=0.16 – 0.18). *Proteobacteria* have previously been associated with gut dysbiosis, though associations are often inconsistent among children from different world regions.^39,40^ *Slackia*, which is not well-described among children but has previously been isolated from both gut and oral infections in adults,^41^ was more abundant at ESBL-negative timepoints though overall abundance was low (0.006%). *Actinobacteria*, which primarily comprise *Bifidobacteria* spp. and which dominate the infant gut,^38^ were also somewhat less abundant at ESBL-negative timepoints (*p*-*val*=0.12). Differences between our study and previous work^35,36^ could be a result of our focus on the pediatric microbiome, differences in our sequencing approach, differences in environmental exposures, or other factors. We note that whether these differentially abundant taxa preceded or followed ESBL-E gut colonization cannot be distinguished through our analytical approach.

Overall, the fact that we did not identify gut taxa or metabolic pathways that were differentially abundant between rarely and frequently colonized children could be because such differences do not exist, or, we may have been underpowered to detect these differences given our modest sample size. Breastfeeding was exceptionally common in this setting and given how strongly maternal breastmilk shapes the infant gut microbiome,^42^ this may have further obscured or minimized gut microbiome differences between groups. Given the high and increasing rates of ESBL-E gut colonization in community settings in LMICs, we note that enrolling sufficient children to identify small effect sizes may be challenging in future studies. Alternatively, other gut microbiome differences that we were unable to explore may underlie differences in ESBL-E gut colonization patterns in this setting, *e.g.*, higher bacteriocin production among commensal strains harbored by rarely colonized children, which could favor the exclusion of ESBL-E.^43^. Interestingly, strain-level analyses using the StrainGE toolkit did identify lower richness of *Klebsiella spp.* and *Enterobacter* spp. among rarely versus frequently colonized children, suggesting differences in gut microbial dynamics could exist at the strain level between these groups. Other studies have found that certain ESBL-E strain/gene combinations are more likely to persist in the gut than others.^44,45^ However, manipulating gut microbial strain dynamics is not likely to be a feasible approach for protecting children from ESBL-E colonization through future interventions.

This study had strengths and limitations. Our study represents the first longitudinal analysis of the gut microbiome of South American children, whose fecal metagenomes are under-represented in global databases.^28^ Because daily surveys were collected, we were able to control for time-varying exposures that are known to affect the gut microbiome, including antibiotic use and recent diarrhea, with a low risk of recall bias. However, our study was relatively small, and we may have been underpowered to detect differences in taxonomic composition and functional potential between groups. Data on maternal antibiotic use during the prenatal period, including intrapartum antibiotic prophylaxis, were not collected for the parent study and likely had important effects on the trajectory of children’s gut microbiome development, but we could not consider those effects here. Further, as previously noted,^4^ we used culture-based methods with a relatively high lower limit of detection (∼2 log10 CFU/g wet feces) to identify ESBL-E gut colonization for this study. Rarely colonized children may have been gut-colonized with ESBL-E at timepoints where they tested negative, just at concentrations below the limit of detection. Since higher concentrations of multidrug-resistant Enterobacterales in the gut are associated with elevated risks of infection,^14,15^ identifying whether specific gut taxa may suppress the growth of ESBL-E or the proliferation of *bla*_CTX-M_ genes remains a clinically relevant question, especially in LMICs where multidrug-resistant infections are becoming increasingly common.^46,47^ Finally, there is no universally accepted definition of rare versus persistent ESBL-E colonization, and studies have employed different definitions.^44,48^ It is possible that using different thresholds for classifying rare versus frequent ESBL-E colonization would yield different results.

## Conclusion

This study examined whether differences in Peruvian children’s gut microbiome development could underpin differences in early-life susceptibility to ESBL-E gut colonization. We did not identify any significant differences in species richness, Shannon diversity, *E. coli* strain diversity, or the relative abundance of specific taxa or metabolic pathways between rarely versus frequently colonized children, suggesting that differences in gut microbiome development are unlikely to explain ESBL-E gut colonization patterns in this setting. Regardless of gut colonization patterns over time, children harbored a lower relative abundance of *Slackia* spp., more unique *E. coli* strains, and significantly more *bla*_CTX-M_ gene copies at timepoints where they were ESBL-E positive. Overall, our findings are in line with recent studies among healthy adults^32–34^ suggesting that differences in the gut microbiome are unlikely to underlie patterns of susceptibility to ESBL-E gut-colonization. Protecting children from ESBL-E gut colonization will likely require that their early-life exposures to ESBL-E reservoirs are minimized.

## Data Availability

Fecal metagenomes are available in NCBI Sequence Read Archive (https://www.ncbi.nlm.nih.gov/sra) under BioProject number PRJNA1138246.

## Acknowledgements

We thank all field workers and the community of Villa El Salvador for their collaboration in the parent study. A preprint of this manuscript was previously posted on medRxiv (doi: https://doi.org/10.1101/2024.11.06.24316595).

## Data Availability

Children’s fecal metagenomes are available in NCBI’s Sequence Read Archive (https://www.ncbi.nlm.nih.gov/sra) under BioProject number PRJNA1138246.

## Ethics approval and consent to participate

The parent cohort study was approved by the Institutional Review Boards of Asociación Benefica PRISMA, Universidad Peruana Cayetano Heredia, and Johns Hopkins University. Written informed consent was obtained from caretakers of the infants for both study participation and the use of collected specimens for subsequent research. The analysis of stool specimens for this substudy was approved by the Institutional Review Boards of Asociación Benefica PRISMA and Universidad Peruana Cayetano Heredia (no. 201592). Metagenomic sequencing of stool specimens was determined to be exempt by the Tufts University Health Sciences IRB (STUDY00001328).

## References

1 Cheikh A, Belefquih B, Chajai Y, Cheikhaoui Y, El Hassani A, Benouda A. Enterobacteriaceae producing extended-spectrum β-lactamases (ESBLs) colonization as a risk factor for developing ESBL infections in pediatric cardiac surgery patients: ‘retrospective cohort study’. BMC Infect Dis 2017; 17: 237.

2 Ben-Ami R, Schwaber MJ, Navon-Venezia S, et al. Influx of extended-spectrum beta-lactamase-producing enterobacteriaceae into the hospital. Clin Infect Dis 2006; 42: 925–34.

3 Denis B, Lafaurie M, Donay J-L, et al. Prevalence, risk factors, and impact on clinical outcome of extended-spectrum beta-lactamase-producing Escherichia coli bacteraemia: a five-year study. Int J Infect Dis 2015; 39: 1–6.

4 Nadimpalli ML, Rojas Salvatierra L, Chakraborty S, et al. Effects of breastfeeding on children’s gut colonization with multidrug-resistant Enterobacterales in peri-urban Lima, Peru. Gut Microbes 2024; 16: 2309681.

5 Nadimpalli ML, Marks SJ, Montealegre MC, et al. Urban informal settlements as hotspots of antimicrobial resistance and the need to curb environmental transmission. Nature Microbiology 2020; 5: 787–95.

6 Singh SR, Mao B, Evdokimov K, et al. Prevalence of MDR organism (MDRO) carriage in children and their household members in Siem Reap Province, Cambodia. JAC-Antimicrobial Resistance 2020; 2. DOI:10.1093/jacamr/dlaa097.

7 Caudell MA, Ayodo C, Ita T, et al. Risk Factors for Colonization With Multidrug-Resistant Bacteria in Urban and Rural Communities in Kenya: An Antimicrobial Resistance in Communities and Hospitals (ARCH) Study. Clin Infect Dis 2023; 77: S104–10.

8 Lautenbach E, Mosepele M, Smith RM, et al. Risk Factors for Community Colonization With Extended-Spectrum Cephalosporin-Resistant Enterobacterales (ESCrE) in Botswana: An Antibiotic Resistance in Communities and Hospitals (ARCH) Study. Clin Infect Dis 2023; 77: S89–96.

9 Araos R, Smith RM, Styczynski A, et al. High Burden of Intestinal Colonization With Antimicrobial-Resistant Bacteria in Chile: An Antibiotic Resistance in Communities and Hospitals (ARCH) Study. Clin Infect Dis 2023; 77: S75–81.

10 Kothari C, Gaind R, Singh LC, et al. Community acquisition of β-lactamase producing Enterobacteriaceae in neonatal gut. BMC Microbiol 2013; 13: 136.

11 Islam MA, Amin MB, Roy S, et al. Fecal colonization with multidrug-resistant E. coli among healthy infants in rural Bangladesh. Frontiers in Microbiology 2019; 10: 640.

12 Tellevik MG, Blomberg B, Kommedal Ø, Maselle SY, Langeland N, Moyo SJ. High Prevalence of Faecal Carriage of ESBL-Producing Enterobacteriaceae among Children in Dar es Salaam, Tanzania. PLOS ONE 2016; 11: e0168024.

13 Lawley TD, Walker AW. Intestinal colonization resistance. Immunology 2013; 138: 1–11.

14 Ruppé E, Lixandru B, Cojocaru R, et al. Relative fecal abundance of extended-spectrum-β-lactamase-producing Escherichia coli strains and their occurrence in urinary tract infections in women. Antimicrob Agents Chemother 2013; 57: 4512–7.

15 Shimasaki T, Seekatz A, Bassis C, et al. Increased Relative Abundance of Klebsiella pneumoniae Carbapenemase-producing Klebsiella pneumoniae Within the Gut Microbiota Is Associated With Risk of Bloodstream Infection in Long-term Acute Care Hospital Patients. Clin Infect Dis 2019; 68: 2053–9.

16 Pajuelo MJ, Noazin S, Cabrera L, et al. Epidemiology of enterotoxigenic Escherichia coli and impact on the growth of children in the first two years of life in Lima, Peru. Front Public Health 2024; 12: 1332319.

17 Stewart CJ, Ajami NJ, O’Brien JL, et al. Temporal development of the gut microbiome in early childhood from the TEDDY study. Nature 2018; 562: 583–8.

18 Brian Bushnell. BBMap. https://sourceforge.net/projects/bbmap/.

19 Truong DT, Tett A, Pasolli E, Huttenhower C, Segata N. Microbial strain-level population structure and genetic diversity from metagenomes. Genome Res 2017; 27: 626–38.

20 Beghini F, McIver LJ, Blanco-Míguez A, et al. Integrating taxonomic, functional, and strain-level profiling of diverse microbial communities with bioBakery 3. Elife 2021; 10: e65088.

21 Langmead B, Salzberg SL. Fast gapped-read alignment with Bowtie 2. Nat Methods 2012; 9: 357–9.

22 Suzek BE, Wang Y, Huang H, McGarvey PB, Wu CH, UniProt Consortium. UniRef clusters: a comprehensive and scalable alternative for improving sequence similarity searches. Bioinformatics 2015; 31: 926–32.

23 Buchfink B, Reuter K, Drost H-G. Sensitive protein alignments at tree-of-life scale using DIAMOND. Nat Methods 2021; 18: 366–8.

24 Caspi R, Billington R, Keseler IM, et al. The MetaCyc database of metabolic pathways and enzymes - a 2019 update. Nucleic Acids Research 2020; 48: D445–53.

25 Ye Y, Doak TG. A Parsimony Approach to Biological Pathway Reconstruction/Inference for Genomes and Metagenomes. PLOS Computational Biology 2009; 5: e1000465.

26 Clausen PTLC, Aarestrup FM, Lund O. Rapid and precise alignment of raw reads against redundant databases with KMA. BMC Bioinformatics 2018; 19: 307.

27 Nayfach S, Pollard KS. Average genome size estimation improves comparative metagenomics and sheds light on the functional ecology of the human microbiome. Genome Biol 2015; 16: 51.

28 Fuhrmeister ER, Harvey AP, Nadimpalli ML, et al. Evaluating the relationship between community water and sanitation access and the global burden of antibiotic resistance: an ecological study. The Lancet Microbe 2023; 4: e591–600.

29 Hendriksen RS, Munk P, Njage P, et al. Global monitoring of antimicrobial resistance based on metagenomics analyses of urban sewage. Nature Communications 2019; 10: 1124.

30 Van Dijk LR, Walker BJ, Straub TJ, et al. StrainGE: a toolkit to track and characterize low-abundance strains in complex microbial communities. Genome Biol 2022; 23: 74.

31 Waters NR, Abram F, Brennan F, Holmes A, Pritchard L. Easy phylotyping of Escherichia coli via the EzClermont web app and command-line tool. Access Microbiol 2020; 2: acmi000143.

32 Worby CJ, Sridhar S, Turbett SE, et al. Gut microbiome perturbation, antibiotic resistance, and Escherichia coli strain dynamics associated with international travel: a metagenomic analysis. The Lancet Microbe 2023; 0. DOI:10.1016/S2666-5247(23)00147-7.

33 Ducarmon QR, Zwittink RD, Willems RPJ, et al. Gut colonisation by extended-spectrum β-lactamase-producing Escherichia coli and its association with the gut microbiome and metabolome in Dutch adults: a matched case-control study. Lancet Microbe 2022; 3: e443– 51.

34 Boyd A, El Dani M, Ajrouche R, et al. Gut microbiome diversity and composition in individuals with and without extended-spectrum β-lactamase-producing *Enterobacterales* carriage: a matched case–control study in infectious diseases department. Clinical Microbiology and Infection 2024; published online March 23. DOI:10.1016/j.cmi.2024.03.016.

35 Gosalbes MJ, Vázquez-Castellanos JF, Angebault C, et al. Carriage of Enterobacteria Producing Extended-Spectrum β-Lactamases and Composition of the Gut Microbiota in an Amerindian Community. Antimicrob Agents Chemother 2016; 60: 507–14.

36 Piewngam P, Quiñones M, Thirakittiwatthana W, Yungyuen T, Otto M, Kiratisin P. Composition of the intestinal microbiota in extended-spectrum β-lactamase-producing Enterobacteriaceae carriers and non-carriers in Thailand. Int J Antimicrob Agents 2019; 53: 435–41.

37 Milani C, Duranti S, Bottacini F, et al. The First Microbial Colonizers of the Human Gut: Composition, Activities, and Health Implications of the Infant Gut Microbiota. Microbiology and Molecular Biology Reviews 2017; 81: 10.1128/mmbr.00036-17.

38 Deering KE, Devine A, O’Sullivan TA, Lo J, Boyce MC, Christophersen CT. Characterizing the Composition of the Pediatric Gut Microbiome: A Systematic Review. Nutrients 2020; 12: 16.

39 Shin N-R, Whon TW, Bae J-W. Proteobacteria: microbial signature of dysbiosis in gut microbiota. Trends Biotechnol 2015; 33: 496–503.

40 Rizzatti G, Lopetuso LR, Gibiino G, Binda C, Gasbarrini A. Proteobacteria: A Common Factor in Human Diseases. Biomed Res Int 2017; 2017: 9351507.

41 Kim K-S, Rowlinson M-C, Bennion R, et al. Characterization of Slackia exigua isolated from human wound infections, including abscesses of intestinal origin. J Clin Microbiol 2010; 48: 1070–5.

42 Gopalakrishna KP, Hand TW. Influence of Maternal Milk on the Neonatal Intestinal Microbiome. Nutrients 2020; 12: 823.

43 Majeed H, Gillor O, Kerr B, Riley MA. Competitive interactions in Escherichia coli populations: the role of bacteriocins. ISME J 2011; 5: 71–81.

44 van Duijkeren E, Wielders CCH, Dierikx CM, et al. Long-term Carriage of Extended-Spectrum β-Lactamase–Producing Escherichia coli and Klebsiella pneumoniae in the General Population in The Netherlands. Clinical Infectious Diseases 2018; 66: 1368–76.

45 Armand-Lefèvre L, Rondinaud E, Desvillechabrol D, et al. Dynamics of extended-spectrum beta-lactamase-producing Enterobacterales colonization in long-term carriers following travel abroad. Microbial Genomics 2021; 7: 000576.

46 Hussein RA, AL-Kubaisy SH, Al-Ouqaili MTS. The influence of efflux pump, outer membrane permeability and β-lactamase production on the resistance profile of multi, extensively and pandrug resistant *Klebsiella pneumoniae*. Journal of Infection and Public Health 2024; 17: 102544.

47 Antimicrobial Resistance Collaborators. Global burden of bacterial antimicrobial resistance in 2019: a systematic analysis. Lancet 2022; 399: 629–55.

48 van den Bunt G, Fluit AC, Bootsma MCJ, et al. Dynamics of Intestinal Carriage of Extended-Spectrum Beta-lactamase–Producing Enterobacteriaceae in the Dutch General Population, 2014–2016. Clinical Infectious Diseases 2020; 71: 1847–55.

