## Supplemental File for "Role of the gut microbiome in frequent gut colonization with extended-spectrum β lactamase-producing Enterobacterales among Peruvian children"

**Appendix**

| **List of Tables** | **Page** |
| --- | --- |
| Table S1: SRA accession numbers for 298 fecal metagenomes from 54 Peruvian children, BioProject PRJNA1138246. | 2-9 |
| Table S2: Exposures to breastfeeding in 30 days prior to stool sampling among 54 Peruvian infants, by age at time of stool sampling. | 10 |
| Table S3: Distributions of sociodemographic characteristics and early-life exposures among Peruvian children participating in the parent cohort, children previously screened for ESBL-E gut colonization, and children whose fecal metagenomes were included in the present study. | 11 |
| Table S4: Potential human pathogens detected in 54 Peruvian children’s stool at 3,6,7,9,12 and 16 months of life. | 12-14 |

| **List of Figures** |  |
| --- | --- |
| Figure S1: Gut colonization patterns among 12 Peruvian children who were rarely colonized with ESBL-producing *E. coli* or ESBL-producing KEC during the first 16 months of life, as identified using culture-based methods | 15 |
| Figure S2: Gut colonization patterns among 42 Peruvian children who were frequently colonized with ESBL-producing *E. coli* or ESBL-producing KEC during the first 16 months of life, as identified using culture-based methods. | 16 |
| Figure S3: Changes in alpha diversity of Peruvian children’s fecal metagenomes from 3-16 months of age, stratified by frequency of gut-colonization with ESBL-producing Enterobacterales during this time frame. | 17 |
| Figure S4: Abundance of A) All antibiotic resistance genes and B) *bla*_CTX-M_ resistance genes among Peruvian children’s fecal metagenomes from 3-16 months of age, stratified by frequency of gut-colonization with ESBL-producing Enterobacterales during this time frame. | 18 |
| Figure S5: Taxa A) positively and B) negatively associated with ESBL-E gut carriage at any given time point. | 19 |

**Table S1.** SRA accession numbers for 298 fecal metagenomes from 54 Peruvian children, BioProject PRJNA1138246.

| Sample | BioSample Accession Number | Child ID | Rounded child age (months) | ESBL-E gut colonization status |
| --- | --- | --- | --- | --- |
| SVHH05013 | SAMN42686833 | 1 | 3 | frequent |
| SVHH08544 | SAMN42686861 | 1 | 6 | frequent |
| SVHH09826 | SAMN42686874 | 1 | 7 | frequent |
| SVHH12235 | SAMN42686918 | 1 | 9 | frequent |
| SVHH16871 | SAMN42687002 | 1 | 12 | frequent |
| SVHH21297 | SAMN42687066 | 1 | 16 | frequent |
| SVHH05399 | SAMN42686834 | 2 | 3 | frequent |
| SVHH09115 | SAMN42686866 | 2 | 6 | frequent |
| SVHH12945 | SAMN42686928 | 2 | 9 | frequent |
| SVHH16642 | SAMN42686998 | 2 | 12 | frequent |
| SVHH21860 | SAMN42687073 | 2 | 16 | frequent |
| SVHH04418 | SAMN42686832 | 3 | 3 | frequent |
| SVHH07977 | SAMN42686854 | 3 | 6 | frequent |
| SVHH10022 | SAMN42686878 | 3 | 7 | frequent |
| SVHH11834 | SAMN42686908 | 3 | 9 | frequent |
| SVHH16731 | SAMN42686999 | 3 | 12 | frequent |
| SVHH20831 | SAMN42687061 | 3 | 16 | frequent |
| SVHH05967 | SAMN42686837 | 4 | 3 | frequent |
| SVHH09607 | SAMN42686870 | 4 | 6 | frequent |
| SVHH11031 | SAMN42686898 | 4 | 7 | frequent |
| SVHH13493 | SAMN42686945 | 4 | 9 | frequent |
| SVHH17158 | SAMN42687007 | 4 | 12 | frequent |
| SVHH22267 | SAMN42687078 | 4 | 16 | frequent |
| SVHH05824 | SAMN42686835 | 5 | 3 | frequent |
| SVHH09497 | SAMN42686869 | 5 | 6 | frequent |
| SVHH10786 | SAMN42686889 | 5 | 7 | frequent |
| SVHH13093 | SAMN42686934 | 5 | 9 | frequent |
| SVHH17439 | SAMN42687015 | 5 | 12 | frequent |
| SVHH21890 | SAMN42687074 | 5 | 16 | frequent |
| SVHH05849 | SAMN42686836 | 6 | 3 | rare |
| SVHH09716 | SAMN42686871 | 6 | 6 | rare |
| SVHH11030 | SAMN42686897 | 6 | 7 | rare |
| SVHH13368 | SAMN42686942 | 6 | 9 | rare |
| SVHH17406 | SAMN42687013 | 6 | 12 | rare |
| SVHH06339 | SAMN42686839 | 7 | 3 | frequent |
| SVHH10023 | SAMN42686879 | 7 | 6 | frequent |
| SVHH11092 | SAMN42686899 | 7 | 7 | frequent |
| SVHH13633 | SAMN42686948 | 7 | 9 | frequent |
| SVHH17530 | SAMN42687016 | 7 | 12 | frequent |
| SVHH22651 | SAMN42687084 | 7 | 16 | frequent |
| SVHH06693 | SAMN42686842 | 8 | 3 | frequent |
| SVHH10864 | SAMN42686890 | 8 | 7 | frequent |
| SVHH17249 | SAMN42687010 | 8 | 12 | frequent |
| SVHH22499 | SAMN42687081 | 8 | 16 | frequent |
| SVHH08486 | SAMN42686860 | 9 | 3 | frequent |
| SVHH11001 | SAMN42686895 | 9 | 6 | frequent |
| SVHH14827 | SAMN42686968 | 9 | 9 | frequent |
| SVHH18668 | SAMN42687028 | 9 | 12 | frequent |
| SVHH23230 | SAMN42687089 | 9 | 16 | frequent |
| SVHH06820 | SAMN42686844 | 10 | 3 | frequent |
| SVHH10583 | SAMN42686886 | 10 | 6 | frequent |
| SVHH12162 | SAMN42686916 | 10 | 7 | frequent |
| SVHH15969 | SAMN42686984 | 10 | 9 | frequent |
| SVHH18546 | SAMN42687026 | 10 | 12 | frequent |
| SVHH23154 | SAMN42687087 | 10 | 16 | frequent |
| SVHH06221 | SAMN42686838 | 11 | 3 | rare |
| SVHH10090 | SAMN42686882 | 11 | 6 | rare |
| SVHH11284 | SAMN42686905 | 11 | 7 | rare |
| SVHH13653 | SAMN42686949 | 11 | 9 | rare |
| SVHH19003 | SAMN42687036 | 11 | 12 | rare |
| SVHH22597 | SAMN42687083 | 11 | 16 | rare |
| SVHH06604 | SAMN42686840 | 12 | 3 | frequent |
| SVHH11802 | SAMN42686907 | 12 | 7 | frequent |
| SVHH14311 | SAMN42686956 | 12 | 9 | frequent |
| SVHH18293 | SAMN42687023 | 12 | 12 | frequent |
| SVHH22859 | SAMN42687085 | 12 | 16 | frequent |
| SVHH06800 | SAMN42686843 | 13 | 3 | frequent |
| SVHH10585 | SAMN42686887 | 13 | 6 | frequent |
| SVHH14689 | SAMN42686965 | 13 | 9 | frequent |
| SVHH18464 | SAMN42687024 | 13 | 12 | frequent |
| SVHH22934 | SAMN42687086 | 13 | 16 | frequent |
| SVHH06638 | SAMN42686841 | 14 | 3 | frequent |
| SVHH11228 | SAMN42686903 | 14 | 6 | frequent |
| SVHH13789 | SAMN42686951 | 14 | 9 | frequent |
| SVHH18642 | SAMN42687027 | 14 | 12 | frequent |
| SVHH22574 | SAMN42687082 | 14 | 16 | frequent |
| SVHH07163 | SAMN42686846 | 15 | 3 | frequent |
| SVHH10850 | SAMN42686891 | 15 | 6 | frequent |
| SVHH14872 | SAMN42686969 | 15 | 9 | frequent |
| SVHH18756 | SAMN42687031 | 15 | 12 | frequent |
| SVHH23307 | SAMN42687090 | 15 | 16 | frequent |
| SVHH07302 | SAMN42686847 | 16 | 3 | frequent |
| SVHH11124 | SAMN42686900 | 16 | 6 | frequent |
| SVHH12648 | SAMN42686925 | 16 | 7 | frequent |
| SVHH15285 | SAMN42686975 | 16 | 9 | frequent |
| SVHH19082 | SAMN42687037 | 16 | 12 | frequent |
| SVHH23452 | SAMN42687093 | 16 | 16 | frequent |
| SVHH07922 | SAMN42686853 | 17 | 3 | frequent |
| SVHH11875 | SAMN42686910 | 17 | 6 | frequent |
| SVHH13041 | SAMN42686932 | 17 | 7 | frequent |
| SVHH16748 | SAMN42687001 | 17 | 9 | frequent |
| SVHH19457 | SAMN42687040 | 17 | 12 | frequent |
| SVHH23731 | SAMN42687098 | 17 | 16 | frequent |
| SVHH08187 | SAMN42686855 | 18 | 3 | frequent |
| SVHH12070 | SAMN42686913 | 18 | 6 | frequent |
| SVHH13258 | SAMN42686940 | 18 | 7 | frequent |
| SVHH14687 | SAMN42686964 | 18 | 9 | frequent |
| SVHH19633 | SAMN42687042 | 18 | 12 | frequent |
| SVHH23426 | SAMN42687092 | 18 | 16 | frequent |
| SVHH07028 | SAMN42686845 | 19 | 3 | frequent |
| SVHH10981 | SAMN42686894 | 19 | 6 | frequent |
| SVHH12294 | SAMN42686919 | 19 | 7 | frequent |
| SVHH14779 | SAMN42686967 | 19 | 9 | frequent |
| SVHH18741 | SAMN42687030 | 19 | 12 | frequent |
| SVHH23586 | SAMN42687096 | 19 | 16 | frequent |
| SVHH07327 | SAMN42686848 | 20 | 3 | frequent |
| SVHH10095 | SAMN42686883 | 20 | 6 | frequent |
| SVHH13791 | SAMN42686955 | 20 | 9 | frequent |
| SVHH17706 | SAMN42687018 | 20 | 12 | frequent |
| SVHH21520 | SAMN42687069 | 20 | 16 | frequent |
| SVHH07921 | SAMN42686852 | 21 | 3 | frequent |
| SVHH11775 | SAMN42686906 | 21 | 6 | frequent |
| SVHH13042 | SAMN42686933 | 21 | 7 | frequent |
| SVHH15416 | SAMN42686978 | 21 | 9 | frequent |
| SVHH19396 | SAMN42687038 | 21 | 12 | frequent |
| SVHH23830 | SAMN42687099 | 21 | 16 | frequent |
| SVHH07823 | SAMN42686851 | 22 | 3 | frequent |
| SVHH11897 | SAMN42686911 | 22 | 6 | frequent |
| SVHH13006 | SAMN42686931 | 22 | 7 | frequent |
| SVHH15470 | SAMN42686979 | 22 | 9 | frequent |
| SVHH20780 | SAMN42687060 | 22 | 12 | frequent |
| SVHH23697 | SAMN42687097 | 22 | 16 | frequent |
| SVHH08625 | SAMN42686864 | 23 | 3 | rare |
| SVHH13469 | SAMN42686944 | 23 | 7 | rare |
| SVHH15950 | SAMN42686986 | 23 | 9 | rare |
| SVHH19973 | SAMN42687050 | 23 | 12 | rare |
| SVHH24175 | SAMN42687105 | 23 | 16 | rare |
| SVHH07775 | SAMN42686850 | 24 | 3 | frequent |
| SVHH11860 | SAMN42686909 | 24 | 6 | frequent |
| SVHH12948 | SAMN42686929 | 24 | 7 | frequent |
| SVHH14543 | SAMN42686961 | 24 | 9 | frequent |
| SVHH20016 | SAMN42687054 | 24 | 12 | frequent |
| SVHH24911 | SAMN42687109 | 24 | 16 | frequent |
| SVHH08605 | SAMN42686863 | 25 | 3 | frequent |
| SVHH15962 | SAMN42686987 | 25 | 9 | frequent |
| SVHH19900 | SAMN42687049 | 25 | 12 | frequent |
| SVHH24126 | SAMN42687104 | 25 | 16 | frequent |
| SVHH08225 | SAMN42686857 | 26 | 3 | frequent |
| SVHH12167 | SAMN42686917 | 26 | 6 | frequent |
| SVHH13179 | SAMN42686937 | 26 | 7 | frequent |
| SVHH15721 | SAMN42686982 | 26 | 9 | frequent |
| SVHH19750 | SAMN42687044 | 26 | 12 | frequent |
| SVHH23953 | SAMN42687100 | 26 | 16 | frequent |
| SVHH08556 | SAMN42686862 | 27 | 3 | frequent |
| SVHH12344 | SAMN42686922 | 27 | 6 | frequent |
| SVHH13614 | SAMN42686947 | 27 | 7 | frequent |
| SVHH16103 | SAMN42686989 | 27 | 9 | frequent |
| SVHH19847 | SAMN42687046 | 27 | 12 | frequent |
| SVHH24262 | SAMN42687107 | 27 | 16 | frequent |
| SVHH07399 | SAMN42686849 | 28 | 3 | frequent |
| SVHH11241 | SAMN42686904 | 28 | 6 | frequent |
| SVHH12754 | SAMN42686927 | 28 | 7 | frequent |
| SVHH15284 | SAMN42686976 | 28 | 9 | frequent |
| SVHH17755 | SAMN42687019 | 28 | 12 | frequent |
| SVHH23550 | SAMN42687094 | 28 | 16 | frequent |
| SVHH08233 | SAMN42686858 | 29 | 3 | rare |
| SVHH12082 | SAMN42686914 | 29 | 6 | rare |
| SVHH13296 | SAMN42686941 | 29 | 7 | rare |
| SVHH17078 | SAMN42687005 | 29 | 9 | rare |
| SVHH19602 | SAMN42687041 | 29 | 12 | rare |
| SVHH25024 | SAMN42687112 | 29 | 16 | rare |
| SVHH09742 | SAMN42686872 | 30 | 3 | frequent |
| SVHH12305 | SAMN42686921 | 30 | 6 | frequent |
| SVHH15935 | SAMN42686985 | 30 | 9 | frequent |
| SVHH19850 | SAMN42687047 | 30 | 12 | frequent |
| SVHH24206 | SAMN42687106 | 30 | 16 | frequent |
| SVHH08937 | SAMN42686865 | 31 | 3 | frequent |
| SVHH12700 | SAMN42686926 | 31 | 6 | frequent |
| SVHH13766 | SAMN42686954 | 31 | 7 | frequent |
| SVHH16469 | SAMN42686995 | 31 | 9 | frequent |
| SVHH20099 | SAMN42687057 | 31 | 12 | frequent |
| SVHH24375 | SAMN42687108 | 31 | 16 | frequent |
| SVHH08333 | SAMN42686859 | 32 | 3 | frequent |
| SVHH12148 | SAMN42686915 | 32 | 6 | frequent |
| SVHH15780 | SAMN42686983 | 32 | 9 | frequent |
| SVHH19723 | SAMN42687043 | 32 | 12 | frequent |
| SVHH23972 | SAMN42687101 | 32 | 16 | frequent |
| SVHH08191 | SAMN42686856 | 33 | 3 | frequent |
| SVHH12465 | SAMN42686924 | 33 | 6 | frequent |
| SVHH16105 | SAMN42686990 | 33 | 9 | frequent |
| SVHH20010 | SAMN42687052 | 33 | 12 | frequent |
| SVHH24042 | SAMN42687103 | 33 | 16 | frequent |
| SVHH09898 | SAMN42686875 | 34 | 3 | frequent |
| SVHH13511 | SAMN42686946 | 34 | 6 | frequent |
| SVHH14915 | SAMN42686971 | 34 | 7 | frequent |
| SVHH17378 | SAMN42687012 | 34 | 9 | frequent |
| SVHH19978 | SAMN42687051 | 34 | 12 | frequent |
| SVHH25414 | SAMN42687118 | 34 | 16 | frequent |
| SVHH09498 | SAMN42686868 | 35 | 3 | frequent |
| SVHH11956 | SAMN42686912 | 35 | 6 | frequent |
| SVHH14471 | SAMN42686962 | 35 | 7 | frequent |
| SVHH17028 | SAMN42687004 | 35 | 9 | frequent |
| SVHH20985 | SAMN42687064 | 35 | 12 | frequent |
| SVHH25104 | SAMN42687114 | 35 | 16 | frequent |
| SVHH09911 | SAMN42686876 | 36 | 3 | frequent |
| SVHH13702 | SAMN42686950 | 36 | 6 | frequent |
| SVHH17795 | SAMN42687020 | 36 | 9 | frequent |
| SVHH20059 | SAMN42687056 | 36 | 12 | frequent |
| SVHH25609 | SAMN42687119 | 36 | 16 | frequent |
| SVHH09806 | SAMN42686873 | 37 | 3 | frequent |
| SVHH13467 | SAMN42686943 | 37 | 6 | frequent |
| SVHH14881 | SAMN42686970 | 37 | 7 | frequent |
| SVHH17231 | SAMN42687009 | 37 | 9 | frequent |
| SVHH22187 | SAMN42687077 | 37 | 12 | frequent |
| SVHH25381 | SAMN42687116 | 37 | 16 | frequent |
| SVHH09322 | SAMN42686867 | 38 | 3 | frequent |
| SVHH13184 | SAMN42686938 | 38 | 6 | frequent |
| SVHH14672 | SAMN42686963 | 38 | 7 | frequent |
| SVHH17198 | SAMN42687008 | 38 | 9 | frequent |
| SVHH20887 | SAMN42687062 | 38 | 12 | frequent |
| SVHH25131 | SAMN42687115 | 38 | 16 | frequent |
| SVHH10857 | SAMN42686892 | 39 | 3 | rare |
| SVHH16024 | SAMN42686988 | 39 | 7 | rare |
| SVHH18887 | SAMN42687033 | 39 | 9 | rare |
| SVHH21353 | SAMN42687068 | 39 | 12 | rare |
| SVHH25408 | SAMN42687117 | 39 | 16 | rare |
| SVHH10482 | SAMN42686885 | 40 | 3 | rare |
| SVHH14406 | SAMN42686960 | 40 | 6 | rare |
| SVHH15316 | SAMN42686977 | 40 | 7 | rare |
| SVHH18041 | SAMN42687021 | 40 | 9 | rare |
| SVHH21315 | SAMN42687067 | 40 | 12 | rare |
| SVHH25711 | SAMN42687120 | 40 | 16 | rare |
| SVHH10727 | SAMN42686888 | 41 | 3 | rare |
| SVHH13142 | SAMN42686935 | 41 | 6 | rare |
| SVHH17134 | SAMN42687006 | 41 | 9 | rare |
| SVHH20911 | SAMN42687063 | 41 | 12 | rare |
| SVHH25026 | SAMN42687113 | 41 | 16 | rare |
| SVHH10061 | SAMN42686877 | 42 | 3 | frequent |
| SVHH13730 | SAMN42686952 | 42 | 6 | frequent |
| SVHH15210 | SAMN42686974 | 42 | 7 | frequent |
| SVHH17572 | SAMN42687017 | 42 | 9 | frequent |
| SVHH21233 | SAMN42687065 | 42 | 12 | frequent |
| SVHH10049 | SAMN42686880 | 43 | 3 | frequent |
| SVHH13008 | SAMN42686930 | 43 | 6 | frequent |
| SVHH14368 | SAMN42686957 | 43 | 7 | frequent |
| SVHH16737 | SAMN42687000 | 43 | 9 | frequent |
| SVHH20659 | SAMN42687059 | 43 | 12 | frequent |
| SVHH24965 | SAMN42687110 | 43 | 16 | frequent |
| SVHH10906 | SAMN42686893 | 44 | 3 | frequent |
| SVHH18726 | SAMN42687029 | 44 | 9 | frequent |
| SVHH21935 | SAMN42687075 | 44 | 12 | frequent |
| SVHH26086 | SAMN42687122 | 44 | 16 | frequent |
| SVHH10087 | SAMN42686881 | 45 | 3 | frequent |
| SVHH13749 | SAMN42686953 | 45 | 6 | frequent |
| SVHH15731 | SAMN42686981 | 45 | 7 | frequent |
| SVHH18997 | SAMN42687035 | 45 | 9 | frequent |
| SVHH21601 | SAMN42687070 | 45 | 12 | frequent |
| SVHH26385 | SAMN42687125 | 45 | 16 | frequent |
| SVHH11011 | SAMN42686896 | 46 | 3 | rare |
| SVHH14758 | SAMN42686966 | 46 | 6 | rare |
| SVHH16144 | SAMN42686991 | 46 | 7 | rare |
| SVHH18718 | SAMN42687032 | 46 | 9 | rare |
| SVHH22306 | SAMN42687079 | 46 | 12 | rare |
| SVHH26118 | SAMN42687123 | 46 | 16 | rare |
| SVHH10284 | SAMN42686884 | 47 | 3 | frequent |
| SVHH14374 | SAMN42686959 | 47 | 6 | frequent |
| SVHH15484 | SAMN42686980 | 47 | 7 | frequent |
| SVHH18167 | SAMN42687022 | 47 | 9 | frequent |
| SVHH21802 | SAMN42687071 | 47 | 12 | frequent |
| SVHH25743 | SAMN42687121 | 47 | 16 | frequent |
| SVHH11134 | SAMN42686901 | 48 | 3 | frequent |
| SVHH15062 | SAMN42686972 | 48 | 6 | frequent |
| SVHH16314 | SAMN42686993 | 48 | 7 | frequent |
| SVHH18980 | SAMN42687034 | 48 | 9 | frequent |
| SVHH22485 | SAMN42687080 | 48 | 12 | frequent |
| SVHH11185 | SAMN42686902 | 49 | 3 | rare |
| SVHH15067 | SAMN42686973 | 49 | 6 | rare |
| SVHH16892 | SAMN42687003 | 49 | 7 | rare |
| SVHH20011 | SAMN42687053 | 49 | 9 | rare |
| SVHH23195 | SAMN42687088 | 49 | 12 | rare |
| SVHH26278 | SAMN42687124 | 49 | 16 | rare |
| SVHH12347 | SAMN42686923 | 50 | 3 | frequent |
| SVHH17311 | SAMN42687011 | 50 | 7 | frequent |
| SVHH19792 | SAMN42687045 | 50 | 9 | frequent |
| SVHH23373 | SAMN42687091 | 50 | 12 | frequent |
| SVHH27104 | SAMN42687126 | 50 | 16 | frequent |
| SVHH13240 | SAMN42686939 | 51 | 3 | rare |
| SVHH16341 | SAMN42686994 | 51 | 6 | rare |
| SVHH20131 | SAMN42687058 | 51 | 9 | rare |
| SVHH24027 | SAMN42687102 | 51 | 12 | rare |
| SVHH13162 | SAMN42686936 | 52 | 3 | frequent |
| SVHH16572 | SAMN42686996 | 52 | 6 | frequent |
| SVHH18492 | SAMN42687025 | 52 | 7 | frequent |
| SVHH20030 | SAMN42687055 | 52 | 9 | frequent |
| SVHH23567 | SAMN42687095 | 52 | 12 | frequent |
| SVHH27615 | SAMN42687128 | 52 | 16 | frequent |
| SVHH12302 | SAMN42686920 | 53 | 3 | rare |
| SVHH16170 | SAMN42686992 | 53 | 6 | rare |
| SVHH17438 | SAMN42687014 | 53 | 7 | rare |
| SVHH19856 | SAMN42687048 | 53 | 9 | rare |
| SVHH22172 | SAMN42687076 | 53 | 12 | rare |
| SVHH27185 | SAMN42687127 | 53 | 16 | rare |
| SVHH14412 | SAMN42686958 | 54 | 3 | rare |
| SVHH16609 | SAMN42686997 | 54 | 6 | rare |
| SVHH19418 | SAMN42687039 | 54 | 7 | rare |
| SVHH21840 | SAMN42687072 | 54 | 9 | rare |
| SVHH24970 | SAMN42687111 | 54 | 12 | rare |
| SVHH28065 | SAMN42687129 | 54 | 16 | rare |

**Table S2.** Exposures to breastfeeding in 30 days prior to stool sampling among 54 Peruvian infants, by age at time of stool sampling.

|  | Proportion of prior 30 days child received breastmilk | | |
| --- | --- | --- | --- |
| Age (months) | Mean | Median | Interquartile Range |
| 3 | 96.6 | 100 | [100,100] |
| 6 | 95.6 | 100 | [100,100] |
| 7 | 94.1 | 100 | [100,100] |
| 9 | 92.7 | 100 | [100,100] |
| 12 | 88.0 | 100 | [100,100] |
| 16 | 82.4 | 100 | [100,100] |

**Table S3**. Distributions of sociodemographic characteristics and early-life exposures among Peruvian children participating in the parent cohort, children previously screened for ESBL-E gut colonization, and children whose fecal metagenomes were included in the present study.

|  | Sequenced for present study | Cohort screened for ESBL-E | Parent cohort who completed 16-month visit |  |
| --- | --- | --- | --- | --- |
|  | (N=54) | (N=112) | (N=274) | *p-val^a^* |
| Infant sex |  |  |  | 0.793 |
| Female | 29 (53.7%) | 57 (50.9%) | 134 (48.9%) |  |
| Male | 25 (46.3%) | 55 (49.1%) | 140 (51.1%) |  |
| Birth mode |  |  |  | 0.030 |
| Cesarean section | 14 (25.9%) | 23 (20.5%) | 35 (12.8%) |  |
| Vaginal | 39 (72.2%) | 78 (69.6%) | 185 (67.5%) |  |
| Missing | 1 (1.9%) | 11 (9.8%) | 54 (19.7%) |  |
| Maternal education |  |  |  | 0.920 |
| At least high school | 34 (63.0%) | 74 (66.1%) | 179 (65.3%) |  |
| Less than high school | 20 (37.0%) | 38 (33.9%) | 94 (34.3%) |  |
| Missing |  |  | 1 (0.4%) |  |
| Type of water source |  |  |  | 0.245 |
| 24-hour indoor  connection | 39 (72.2%) | 87 (77.7%) | 214 (78.1%) |  |
| Public shared tap | 12 (22.2%) | 20 (17.9%) | 36 (13.1%) |  |
| Other | 3 (5.6%) | 5 (4.5%) | 24 (8.8%) |  |
| Toilet type |  |  |  | 0.418 |
| Indoor flush toilet | 42 (77.8%) | 91 (81.3%) | 227 (82.8%) |  |
| Pit latrine | 12 (22.2%) | 19 (17.0%) | 39 (14.2%) |  |
| Other | 0 (0%) | 2 (1.8%) | 8 (2.9%) |  |
| Chickens in the household | |  |  | 0.775 |
| No | 43 (79.6%) | 94 (83.9%) | 228 (83.2%) |  |
| Yes | 11 (20.4%) | 18 (16.1%) | 46 (16.8%) |  |
| Household has improved flooring | |  |  | 0.820 |
| No | 3 (5.6%) | 4 (3.6%) | 13 (4.7%) |  |
| Yes | 51 (94.4%) | 108 (96.4%) | 261 (95.3%) |  |
| Lives in household with more than 6 household members | | | | 0.910 |
| No | 34 (63.0%) | 70 (62.5%) | 166 (60.6%) |  |
| Yes | 20 (37.0%) | 42 (37.5%) | 108 (39.4%) |  |
| Received breastmilk 90% of days in first 3 months | | | | 0.543 |
| No | 22 (40.7%) | 41 (36.6%) | 117 (42.7%) |  |
| Yes | 32 (59.3%) | 71 (63.4) | 157 (57.3%) |  |
| Received formula 50% of days in first 3 months | | |  | 0.777 |
| No | 38 (70.4%) | 84 (75.0%) | 205 (74.8%) |  |
| Yes | 16 (29.6%) | 28 (25.0%) | 69 (25.2%) |  |
| Total number of antibiotic courses per child-year (mean (SD)) | | | | 0.665 |
|  | 4.51 (2.55) | 4.34 (2.82) | 4.18 (2.64) |  |
| Number of diarrhea episodes/100-child-month (mean (SD)) | | | | 0.390 |
|  | 8.12 (8.51) | 8.65 (8.05) | 9.91 (11.92) |  |

^a^Improved flooring included cement, wood, tiles, and “falso piso.” Unimproved flooring included dirt and sand.

**Table S4.** Potential human pathogens detected in 54 Peruvian children’s stool at 3,6,7,9,12 and 16 months of life.

|  |  | No. children colonized at least once | No. samples positive |
| --- | --- | --- | --- |
| Species | Mean relative abundance (%) | N=54 (%) | N=298 (%) |
| *Acinetobacter baumannii* | 2.86E-05 | 1 (2) | 1 (0) |
| *Acinetobacter guillouiae* | 5.45E-05 | 1 (2) | 1 (0) |
| *Acinetobacter haemolyticus* | 6.72E-06 | 1 (2) | 1 (0) |
| *Acinetobacter johnsonii* | 2.32E-05 | 1 (2) | 1 (0) |
| *Acinetobacter ursingii* | 1.34E-04 | 2 (4) | 2 (1) |
| *Aeromonas caviae* | 5.40E-06 | 2 (4) | 2 (1) |
| *Bacteroides caccae* | 4.59E-02 | 25 (46) | 43 (14) |
| *Bacteroides eggerthii* | 1.78E-03 | 9 (17) | 10 (3) |
| *Bacteroides fragilis* | 5.73E-01 | 51 (94) | 171 (57) |
| *Bacteroides galacturonicus* | 5.85E-05 | 3 (6) | 3 (1) |
| *Bacteroides nordii* | 8.88E-04 | 3 (6) | 5 (2) |
| *Bacteroides ovatus* | 4.43E-02 | 38 (70) | 97 (33) |
| *Bacteroides pectinophilus* | 2.96E-03 | 2 (4) | 2 (1) |
| *Bacteroides stercoris* | 9.78E-02 | 25 (46) | 44 (15) |
| *Bacteroides thetaiotaomicron* | 5.73E-02 | 37 (69) | 91 (31) |
| *Bacteroides uniformis* | 4.51E-01 | 50 (93) | 159 (53) |
| *Bacteroides vulgatus* | 2.64E-01 | 46 (85) | 135 (45) |
| *Campylobacter coli* | 6.18E-05 | 2 (4) | 2 (1) |
| *Campylobacter concisus* | 4.31E-04 | 7 (13) | 7 (2) |
| *Campylobacter fetus* | 2.17E-03 | 1 (2) | 1 (0) |
| *Campylobacter jejuni* | 2.68E-02 | 20 (37) | 24 (8) |
| *Clostridium aldenense* | 1.30E-03 | 27 (50) | 45 (15) |
| *Clostridium bolteae* | 4.02E-02 | 32 (59) | 73 (24) |
| *Clostridium butyricum* | 1.75E-05 | 1 (2) | 1 (0) |
| *Clostridium citroniae* | 1.19E-03 | 24 (44) | 40 (13) |
| *Clostridium innocuum* | 2.18E-01 | 53 (98) | 188 (63) |
| *Clostridium neonatale* | 1.72E-03 | 11 (20) | 14 (5) |
| *Clostridium perfringens* | 2.79E-03 | 21 (39) | 27 (9) |
| *Clostridium ventriculi* | 1.73E-04 | 4 (7) | 5 (2) |
| *Corynebacterium accolens* | 1.92E-03 | 21 (39) | 26 (9) |
| *Corynebacterium amycolatum* | 1.22E-02 | 9 (17) | 9 (3) |
| *Corynebacterium argentoratense* | 5.50E-05 | 1 (2) | 1 (0) |
| *Corynebacterium aurimucosum* | 1.61E-04 | 7 (13) | 10 (3) |
| *Corynebacterium durum* | 1.05E-04 | 14 (26) | 15 (5) |
| *Corynebacterium falsenii* | 2.55E-04 | 3 (6) | 3 (1) |
| *Corynebacterium jeikeium* | 2.59E-05 | 3 (6) | 3 (1) |
| *Corynebacterium kroppenstedtii* | 3.15E-04 | 6 (11) | 7 (2) |
| *Corynebacterium riegelii* | 5.08E-06 | 1 (2) | 1 (0) |
| *Corynebacterium simulans* | 1.12E-04 | 3 (6) | 3 (1) |
| *Corynebacterium striatum* | 6.71E-05 | 1 (2) | 1 (0) |
| *Corynebacterium tuberculostearicum* | 1.06E-06 | 1 (2) | 1 (0) |
| *Corynebacterium urealyticum* | 1.28E-05 | 1 (2) | 2 (1) |
| *Enterobacter bugandensis* | 1.13E-05 | 1 (2) | 1 (0) |
| *Enterococcus avium* | 9.14E-02 | 52 (96) | 165 (55) |
| *Enterococcus casseliflavus* | 4.83E-03 | 15 (28) | 20 (7) |
| *Enterococcus cecorum* | 1.09E-02 | 3 (6) | 4 (1) |
| *Enterococcus durans* | 1.21E-02 | 15 (28) | 20 (7) |
| *Enterococcus faecalis* | 3.12E-02 | 42 (78) | 85 (29) |
| *Enterococcus faecium* | 3.87E-01 | 52 (96) | 174 (58) |
| *Enterococcus gallinarum* | 6.39E-02 | 43 (80) | 95 (32) |
| *Enterococcus hirae* | 3.38E-03 | 11 (20) | 11 (4) |
| *Enterococcus mundtii* | 1.75E-05 | 1 (2) | 1 (0) |
| *Enterococcus raffinosus* | 5.69E-03 | 23 (43) | 35 (12) |
| *Helicobacter bilis* | 2.39E-06 | 1 (2) | 1 (0) |
| *Helicobacter cinaedi* | 1.33E-03 | 1 (2) | 1 (0) |
| *Klebsiella aerogenes* | 3.15E-04 | 6 (11) | 9 (3) |
| *Klebsiella oxytoca* | 1.45E-02 | 20 (37) | 27 (9) |
| *Klebsiella pneumoniae* | 5.51E-01 | 50 (93) | 162 (54) |
| *Klebsiella quasipneumoniae* | 7.01E-02 | 47 (87) | 121 (41) |
| *Klebsiella variicola* | 1.31E-01 | 47 (87) | 124 (42) |
| *Neisseria flavescens* | 3.79E-03 | 6 (11) | 7 (2) |
| *Neisseria macacae* | 1.06E-05 | 3 (6) | 3 (1) |
| *Neisseria sicca* | 4.51E-06 | 1 (2) | 1 (0) |
| *Neisseria subflava* | 3.88E-05 | 4 (7) | 4 (1) |
| *Prevotella bivia* | 1.18E-04 | 4 (7) | 4 (1) |
| *Prevotella buccae* | 3.57E-03 | 7 (13) | 10 (3) |
| *Prevotella buccalis* | 1.18E-04 | 5 (9) | 6 (2) |
| *Prevotella corporis* | 3.58E-04 | 7 (13) | 8 (3) |
| *Prevotella dentalis* | 8.90E-06 | 1 (2) | 1 (0) |
| *Prevotella disiens* | 1.98E-05 | 1 (2) | 1 (0) |
| *Prevotella intermedia* | 1.76E-05 | 1 (2) | 1 (0) |
| *Prevotella nigrescens* | 4.50E-04 | 2 (4) | 2 (1) |
| *Prevotella oralis* | 2.60E-04 | 1 (2) | 1 (0) |
| *Prevotella oris* | 8.65E-06 | 1 (2) | 1 (0) |
| *Prevotella veroralis* | 5.23E-06 | 1 (2) | 1 (0) |
| *Salmonella enterica* | 5.72E-05 | 2 (4) | 2 (1) |
| *Staphylococcus aureus* | 5.91E-04 | 15 (28) | 20 (7) |
| *Staphylococcus epidermidis* | 2.21E-03 | 18 (33) | 21 (7) |
| *Staphylococcus haemolyticus* | 2.50E-03 | 26 (48) | 36 (12) |
| *Staphylococcus hominis* | 5.43E-04 | 10 (19) | 13 (4) |
| *Staphylococcus lugdunensis* | 2.37E-05 | 2 (4) | 2 (1) |
| *Streptococcus agalactiae* | 1.07E-05 | 1 (2) | 1 (0) |
| *Streptococcus cristatus* | 1.20E-05 | 1 (2) | 1 (0) |
| *Streptococcus gordonii* | 1.69E-04 | 10 (19) | 11 (4) |
| *Streptococcus infantarius* | 6.93E-03 | 5 (9) | 10 (3) |
| *Streptococcus mitis* | 3.78E-02 | 54 (100) | 247 (83) |
| *Streptococcus mutans* | 1.45E-03 | 9 (17) | 12 (4) |
| *Streptococcus oralis* | 1.10E-03 | 26 (48) | 40 (13) |
| *Streptococcus parasanguinis* | 3.31E-02 | 54 (100) | 261 (88) |
| *Streptococcus pneumoniae* | 4.48E-05 | 6 (11) | 6 (2) |
| *Streptococcus pseudopneumoniae* | 6.33E-04 | 14 (26) | 18 (6) |
| *Streptococcus pyogenes* | 1.17E-05 | 1 (2) | 1 (0) |
| *Streptococcus salivarius* | 1.92E-01 | 54 (100) | 283 (95) |
| *Streptococcus sobrinus* | 2.86E-04 | 7 (13) | 8 (3) |
| *Streptococcus vestibularis* | 2.80E-04 | 18 (33) | 29 (10) |

Note: We examined human pathogens as reported by Bartlett *et al.* 2022 (doi: 10.1099/mic.0.001269). We focused on “established” human pathogens, *i.e*., pathogens that have infected at least three persons in three or more references identified in their review.

Figures

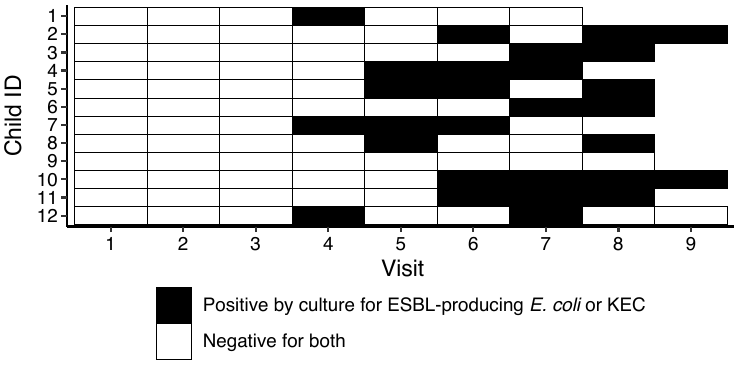

**Figure S1. Gut colonization patterns among 12 Peruvian children who were *rarely* colonized with ESBL-producing *E. coli* or ESBL-producing KEC during the first 16 months of life, as identified using culture-based methods.** Stool samples (visits) were screened at approximately 1, 3, 4, 5, 6, 7, 9, 12, and 16 months of age, but the exact age at which each sample was taken varied among children. Children colonized at visit 4 (IDs 1, 7, and 12) were aged 7.5 months, 6.2 months, and 5.5 months at this timepoint, respectively.

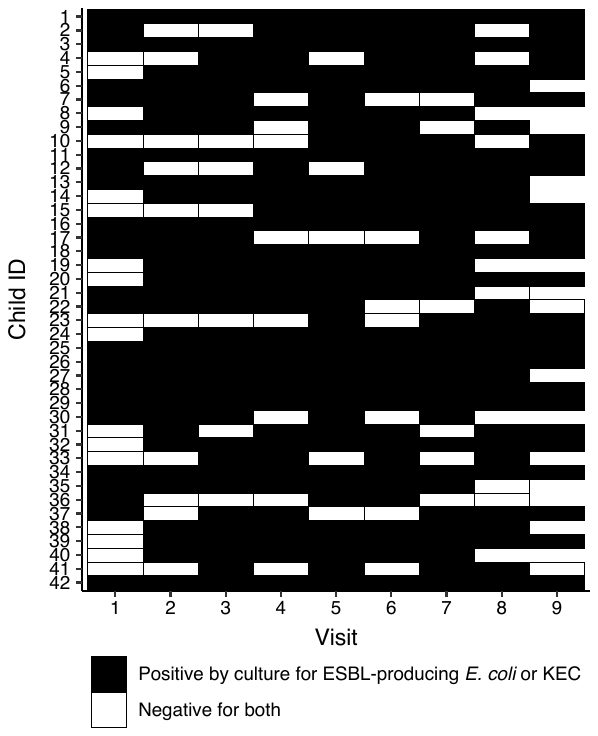

**Figure S2.** Gut colonization patterns among 42 Peruvian children who were *frequently* colonized with ESBL-producing *E. coli* or ESBL-producing KEC during the first 16 months of life, as identified using culture-based methods. Stool samples (visits) were screened at approximately 1, 3, 4, 5, 6, 7, 9, 12, and 16 months of age, but the exact age at which each sample was taken varied among children.

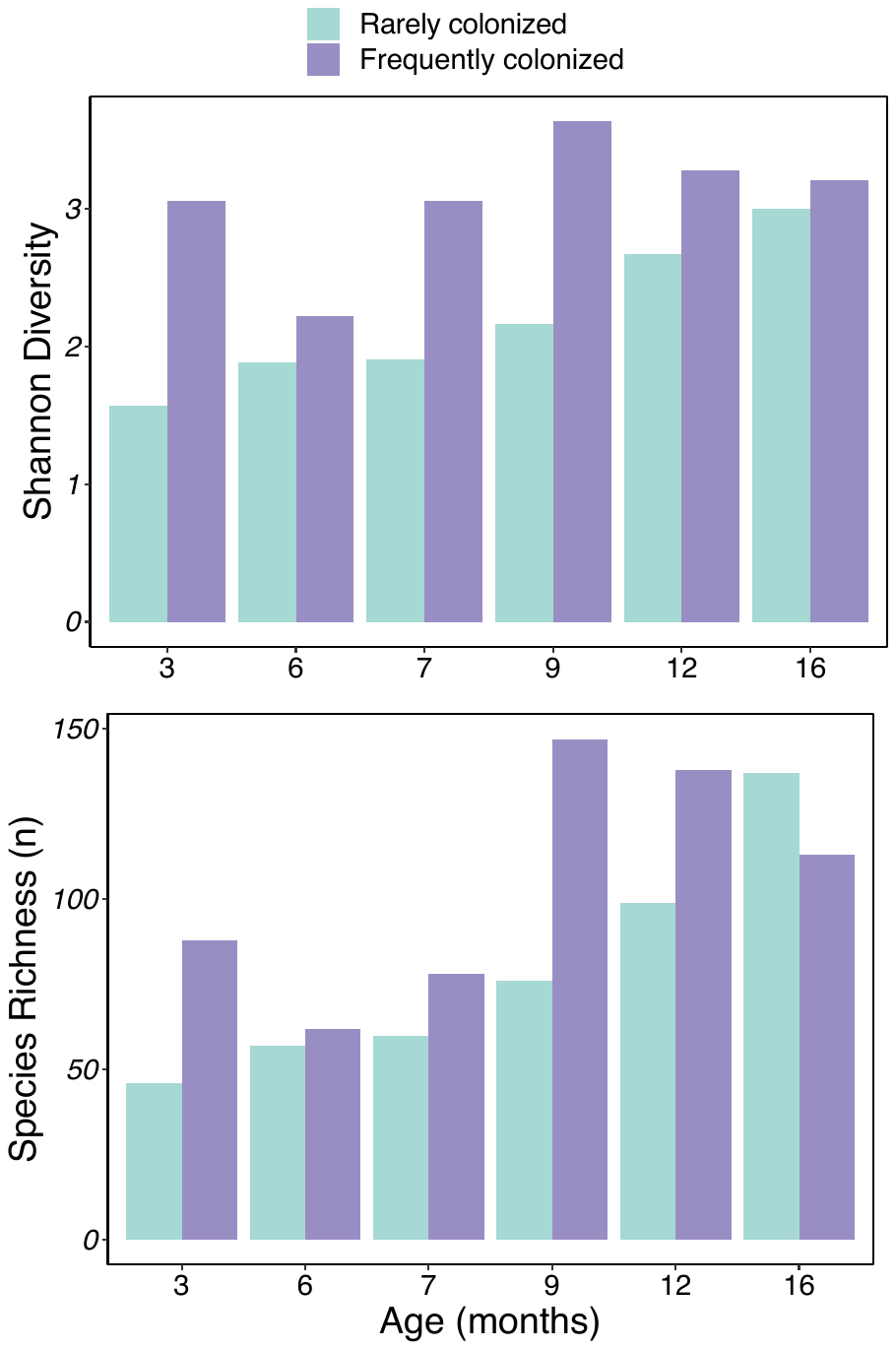

**Figure S3.** Changes in alpha diversity of Peruvian children’s fecal metagenomes from 3-16 months of age, stratified by frequency of gut-colonization with ESBL-producing Enterobacterales during this time frame.

Note**:** Pooling all timepoints together, we observed no differences in the diversity or number of unique species in rarely versus frequently gut-colonized children over time (p-values> 0.62). There were also no differences in median richness or Shannon diversity for any specific age group (p>0.52 for all pairwise differences by Wilcoxon rank sum tests).

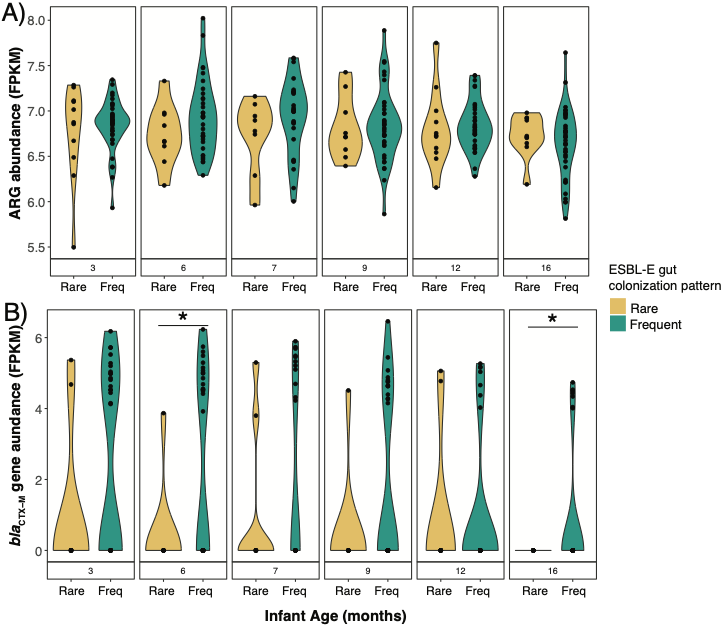
**Figure S4**. Abundance of A) total antibiotic resistance genes and B) *bla*_CTX-M_ resistance genes in Peruvian children’s stool from 3-16 months of age, stratified by frequency of gut-colonization with ESBL-producing Enterobacterales during this time frame. There was no difference in A) total ARG abundance, on average across all timepoints, between rarely and persistently colonized children. The abundance of B) *bla*_CTX-M_ genes was significantly lower among rarely colonized children on average and across all timepoints (*p*<0.0001).), and specifically at ages 6 (*p*=0.002) and 16 months (*p*=0.006) by Welch two-sample test.

*Note*: * denotes a statistically significant difference by Welch two sample t-test. FPKM=Fragments per kilobase per million mapped reads. FPKM values have been log transformed.

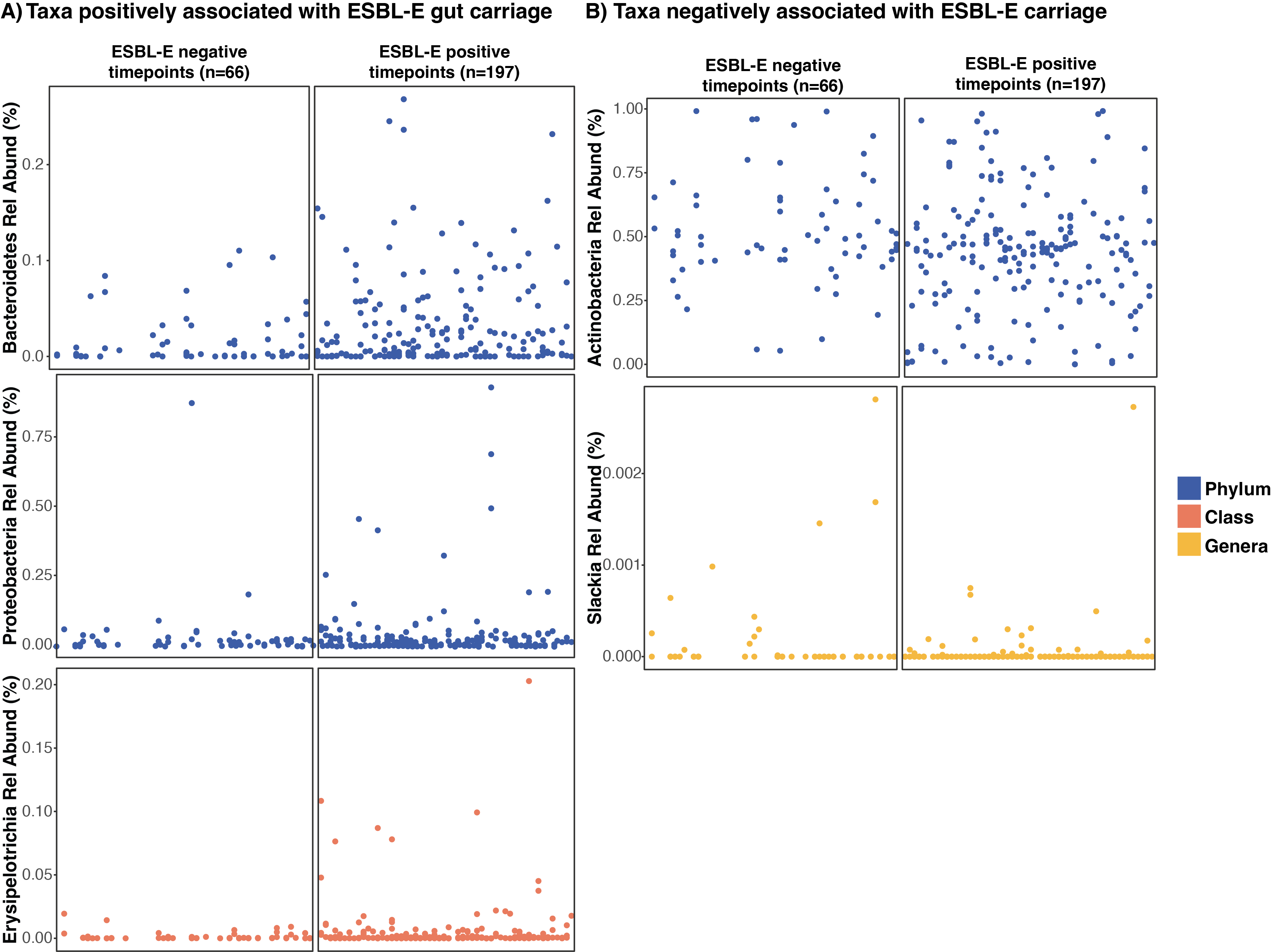

**Figure S5.** Taxa A) positively and B) negatively associated with ESBL-E gut carriage at any given time point.

*Note*: For any given taxa detected in at least 10% of samples, we modeled ESBL-E gut colonization status (yes/no) as a function of subject (random effects term), child age in months, hours between child defecation and field worker retrieval of a child’s diaper, and the transformed relative abundance value for that taxa, using mixed effects logistic regression models.
